# Mitochondrial MICOS complex genes, implicated in hypoplastic left heart syndrome, maintain cardiac contractility and actomyosin integrity

**DOI:** 10.1101/2022.06.13.22276366

**Authors:** Katja Birker, Natalie J. Kirkland, Jeanne L. Theis, Zachary C. Fogarty, Maria Azzurra Missinato, Sreehari Kalvakuri, Paul Grossfeld, Adam J. Engler, Karen Ocorr, Timothy J. Nelson, Alexandre R. Colas, Timothy M. Olson, Georg Vogler, Rolf Bodmer

## Abstract

Hypoplastic left heart syndrome (HLHS) is a severe congenital heart disease (CHD) with a likely oligogenic etiology, but our understanding of the genetic complexities and pathogenic mechanisms leading to HLHS is limited. We therefore performed whole genome sequencing (WGS) on a large cohort of HLHS patients and their families to identify candidate genes that were then tested in *Drosophila* heart model for functional and structural requirements. Bioinformatic analysis of WGS data from an index family comprised of a HLHS proband born to consanguineous parents and postulated to have a homozygous recessive disease etiology, prioritized 9 candidate genes with rare, predicted damaging homozygous variants. Of the candidate HLHS gene homologs tested, cardiac-specific knockdown (KD) of mitochondrial MICOS complex subunit *dCHCHD3/6* resulted in drastically compromised heart contractility, diminished levels of sarcomeric actin and myosin, reduced cardiac ATP levels, and mitochondrial fission-fusion defects. Interestingly, these heart defects were similar to those inflicted by cardiac KD of ATP synthase subunits of the electron transport chain (ETC), consistent with the MICOS complex’s role in maintaining cristae morphology and ETC complex assembly. Analysis of 183 genomes of HLHS patient-parent trios revealed five additional HLHS probands with rare, predicted damaging variants in *CHCHD3* or *CHCHD6*. Hypothesizing an oligogenic basis for HLHS, we tested 60 additional prioritized candidate genes in these cases for genetic interactions with *CHCHD3/6* in sensitized fly hearts. Moderate KD of *CHCHD3/6* in combination with *Cdk12* (activator of RNA polymerase II), *RNF149* (E3 ubiquitin ligase), or *SPTBN1* (scaffolding protein) caused synergistic heart defects, suggesting the potential involvement of a diverse set of pathways in HLHS. Further elucidation of novel candidate genes and genetic interactions of potentially disease-contributing pathways is expected to lead to a better understanding of HLHS and other CHDs.

## INTRODUCTION

Hypoplastic left heart syndrome (HLHS) is a birth defect that accounts for 2-4% of congenital heart defects (CHDs), equal to 1000-2000 HLHS births in the United States per year. HLHS has been proposed to be caused by genetic, epigenetic, or environmental factors (Crucean *et al*., 2017; Liu *et al*., 2017; Yagi *et al*., 2018; Grossfeld *et al*., 2019). The severe cardiac characteristics of HLHS include aortic and mitral stenosis or atresia, and reduced size of the left ventricle and aorta; however, there is a spectrum of cardiac phenotypes that can underly HLHS pathophysiology (Theis, Hrstka, *et al*., 2015; Crucean *et al*., 2017; Mussa and Barron, 2017; Grossfeld *et al*., 2019). If not treated with reconstructive heart surgeries or cardiac transplantation, infants born with HLHS will not survive (Grossfeld *et al*., 2019). To date, the standard treatment for this disease is a three-stage surgical procedure, which begins neonatally and aims overall to achieve right ventricle-dependent systemic circulation and deliver oxygen-poor blood more directly to the lungs (Mussa and Barron, 2017). Although the surgical procedures correctly divert left ventricular function to the right ventricle, there is a subgroup of HLHS patients who are at risk of latent heart failure, which is often preceded by reduced ejection fraction (Altmann *et al*., 2000; Mcbride *et al*., 2008; Theis, Zimmermann, *et al*., 2015).

Although several studies have examined the molecular underpinnings of HLHS, the number of genes associated with this disease is small (e.g. *NKX2-5, NOTCH1, ETS1, MYH6, LRP2* and *CELSR1*), and they are not yet conclusively determined as causal for HLHS (Garg *et al*., 2005; Ye *et al*., 2009; Kobayashi *et al*., 2014; Theis, Hrstka, *et al*., 2015; Tomita-mitchell *et al*., 2016; Theis *et al*., 2020, 2021, 2022). Defining pathogenic mechanisms has proved elusive given the oligogenic complexity of HLHS. Overall, there is a great need to functionally evaluate newly emerging HLHS candidate genes to understand how they may contribute to the molecular, cellular, and morphological processes underlying HLHS.

*Drosophila* is well-suited for modeling genetic underpinnings of CHDs: many of the genes and gene programs found in the *Drosophila* heart are evolutionarily conserved, including a core set of cardiogenic transcription factors and inductive factors (e.g. *Nkx2-5*/*tinman*) (Bodmer, 1995; Cripps and Olson, 2002; Bier and Bodmer, 2004; Bodmer and Frasch, 2010; Ahmad, 2017), approximately 75% of known human disease-causing genes having fly orthologs (Bodmer and Frasch, 2010; Pandey and Nichols, 2011; Ugur, Chen and Bellen, 2016), and the developing mammalian and *Drosophila* hearts share developmental similarities, such as their origin within the mesoderm.

Mitochondria have been postulated to play a critical role in HLHS pathogenesis. For example, a recent study reported that cardiomyocytes derived from iPSCs of HLHS patients (iPSC-CM), who later developed right ventricular failure, had reduced mitochondrial concentration, ATP production, and contractile force (Paige *et al*., 2020). This study revealed downregulated expression of genes involved in mitochondrial processes, such as ATP synthesis coupled electron transport. Another study of HLHS patient-derived iPSC-CMs revealed reduced mitochondrial size, number, and malformed mitochondrial inner membranes using transmission electron microscopy (Yang *et al*., 2017). Similarly, an HLHS mouse model with *Sap130* and *Pcdha9* mutations showed mitochondrial defects manifested as reduced cristae density and smaller mitochondrial size (Liu *et al*., 2017). Despite a lack of understanding of the exact mitochondrial mechanisms underlying HLHS pathogenesis, recent experimental and bioinformatic data suggest an underlying role of mitochondria in HLHS.

Here, a cohort of 183 HLHS proband-parent trios underwent whole genome sequencing (WGS) to identify candidate genes, including a prioritized consanguineous family where genes harboring rare, predicted damaging homozygous variants were investigated (Theis and Olson, 2022). Among the resulting candidate HLHS genes tested in *Drosophila*, cardiac-specific knockdown (KD) of *CHCHD3/6* (*coiled-coil-helix-coiled-coil-helix-domain-containing protein* 6) of the MICOS (mitochondrial contact site and cristae organization system) complex exhibited severe heart structure and function defects. The MICOS complex is an eight-subunit complex in mammals (five in *Drosophila*) located in the inner mitochondrial membrane that is necessary to maintain cristae morphology and ATP production. It is closely associated and interacts with SAM50 (sorting and assembly machinery), which is located in the outer mitochondrial membrane (Ott *et al*., 2012; Kozjak-Pavlovic, 2017). The MICOS complex’s role in cardiac development and functional homeostasis is not known but is likely important for efficient ATP production. We observed reduced contractility upon cardiac-specific *dCHCHD3/6* KD, diminished sarcomeric Actin and Myosin levels, as well as severe mitochondrial morphology defects, which manifested as fragmented and aggregated structures. Similar phenotypes were observed upon cardiac KD of other MICOS complex genes, as well as other mitochondrial genes such as ATP synthase (complex V), specifically ATP synthase B and β. We also found significantly diminished proliferation of human induced pluripotent stem cell (iPSC)-derived ventricular-like cardiomyocytes (VCMs) upon KD of MICOS genes. Finally, a family-based candidate gene interaction screen in *Drosophila* revealed three genes that genetically interact with *dCHCHD3/6*: *Cdk12* (activator RNA polymerase II activator), *RNF149* (E3 ubiquitin ligase), *SPTBN1* (scaffolding protein). In summary, *CHCHD3/6* and other components important for mitochondrial homeostasis were identified as critical for establishing and maintaining cardiac structure and function, and likely contribute to HLHS and/or latent heart failure following surgical palliation.

## RESULTS

### Family Phenotype

Family 11H is of white ancestry and comprised of a male with HLHS, his parents, and two siblings, they were all phenotypically characterized by echocardiography and underwent WGS. A homozygous recessive disease mode of inheritance was postulated due to reported consanguinity between the mother and father and absence of structural and myopathic heart disease in the parents. The siblings also had normal echocardiograms (**Figure 1A**). The 11H proband had latent decline of right ventricular ejection fraction several years after surgical palliation. In addition to HLHS, he was diagnosed with developmental delay, cerebral and cerebellar atrophy, white matter loss, decreased muscle mass, and a body mass index <1%, traits that have previously been related to mitochondrial dysfunction (Romanello and Sandri, 2016; Alston *et al*., 2017).

**Figure 1:**
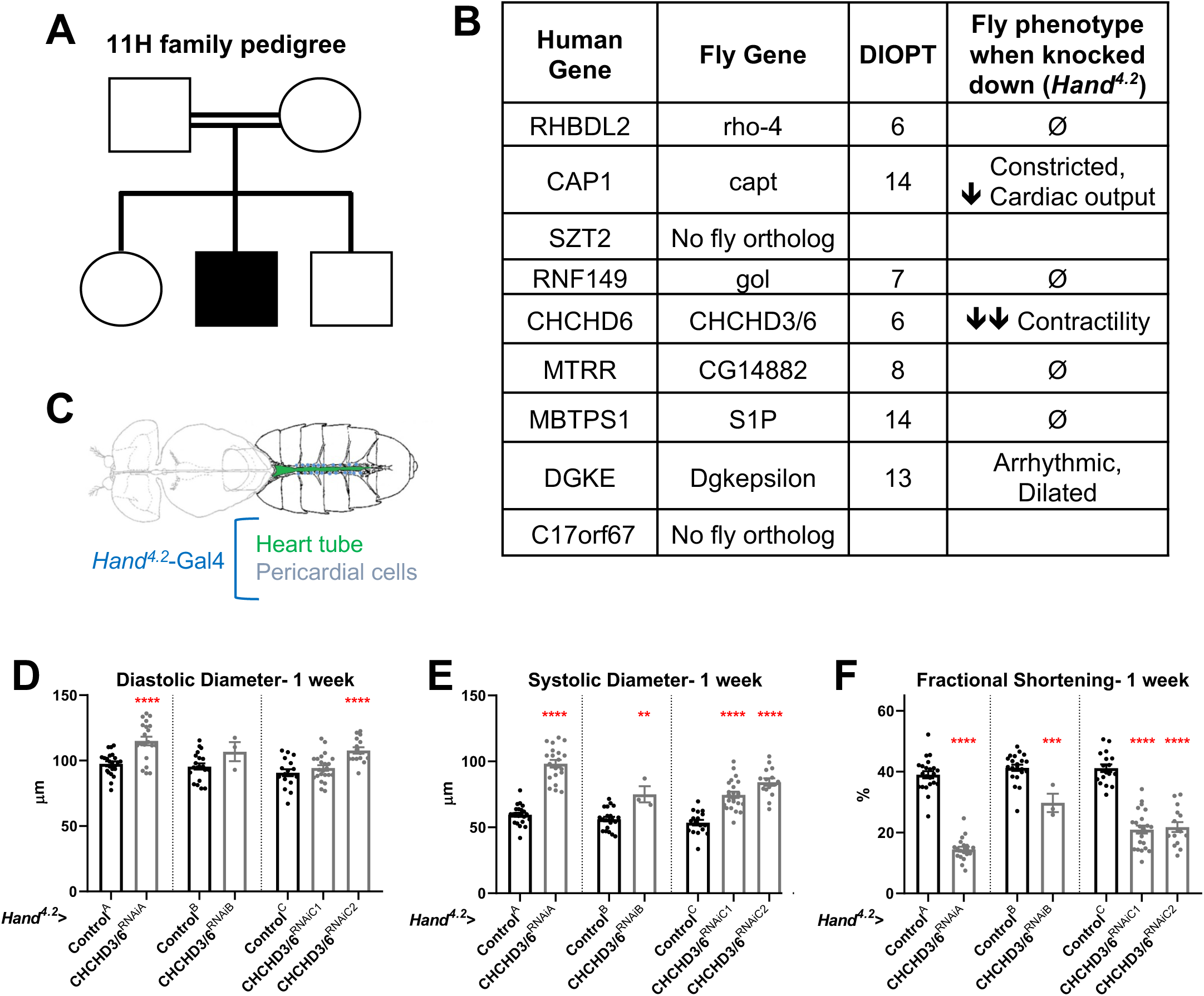
Prioritization of *CHCHD6* in HLHS proband and its *Drosophila* ortholog *CHCHD3/6*. **A)** Pedigree of index family 11H. The family includes consanguineous parents (denoted by double horizontal lines) without cardiac defects, one son with HLHS (proband), and two siblings without cardiac defects. **B)** List of 9 candidate genes derived from proband 11H with corresponding *Drosophila* orthologs. Orthology based on DIOPT score. Conserved *Drosophila* candidate HLHS genes were knocked down individually in the *Drosophila* heart using the *Hand^4.2^-Gal4* driver. The functional phenotypes listed were significantly different relative to Control^A^ or Control^B^ and were measured in 1-week old female *Drosophila* hearts. **C)** Schematic of *Drosophila* highlighting the abdominal region which includes the heart tube and flanking pericardial cells, where the *Hand^4.2^*-Gal4 driver is expressed. Image adapted from (Xie *et al*., 2013). **D)** End-Diastolic diameter (EDD), **E)** End-systolic diameter (ESD), and **F)** fractional shortening (FS) from 1-week old female *Hand^4.2^-Gal4* >*CHCHD3/6* flies.

### Whole Genome Sequencing and Bioinformatics Analysis of 11H family

Array comparative genomic hybridization ruled out a chromosomal deletion or duplication in the proband. WGS was carried out on genomic DNA samples from the five family members, based on paired-end reads that passed quality control standards; 99.4% of the reads mapped to the genome. After marking and filtering out duplicate reads, over 91% of the hg38 human reference genome had coverage. The average depth across the genome was 63X and an average of 89% of the genome demonstrated a minimal read depth of 20 reads. Filtering for rare variants that were homozygous in the HLHS proband revealed nine candidate genes. Three genes had a missense variant (*SZT2*, *MTRR*, *MBTPS1*) whereas the remaining six genes were found to have a non-coding variant within the promoter (*CAP1*, *DGKE*), 5’ untranslated region (*RHBDL2*, *RNF149*, *C17orf67*), or intron (*CHCHD6*) (**Supplementary Table 1**) (Marian and Belmont, 2011). While six of the variants were also found to be homozygous in an unaffected sibling, the associated candidate genes were not excluded from downstream analyses based on the postulated oligogenic nature of HLHS, and incomplete penetrance of individual variants, as observed in a digenic mouse model (Yagi *et al*., 2018).

### Candidate HLHS gene knockdown in *Drosophila* reveals requirement for *CHCHD3/6* in establishing cardiac structure and function

To test whether the HLHS candidates had significant requirements in the heart, we utilized the established *Drosophila* heart model and cardiac-specific RNAi KD. First, the nine candidate genes were assigned their respective *Drosophila* homologs; seven out of nine of the human HLHS candidate genes had *Drosophila* orthologs (**Figure 1B**) (Hu *et al*., 2011). The *Drosophila* Gal4-UAS system (Brand and Perrimon, 1993) was used to test candidate genes for their role in heart function using temporal and/or spatial KD via RNAi. The *Hand^4.2^*-Gal4 driver was used for initial screening because it is a strong post-mitotic and heart-specific driver, which is expressed throughout life in the cardiomyocytes (CMs) and pericardial cells (PCs) (Han and Olson, 2005; Han *et al*., 2006) (**Figure 1C**). 3-week-old (mid-adult stage) female flies were used to test the seven candidate genes. *Hand^4.2^*-Gal4 KD of *capt* (actin binding protein, negatively regulating actin filament assembly)*, Dgkepsilon* (Diacyl glycerol kinase, DGKE), and *CHCHD3/6* (Mitochondrial inner membrane protein of the MICOS complex, required for fusion) produced defects in the fly hearts, such as reduced cardiac output, reduced fractional shortening, and arrhythmicity (**Figure 1B**). Of those, *CHCHD3/6* KD gave the most severe cardiac defects with strongly reduced fractional shortening, a measure of cardiac contractility. Systolic rather than end-diastolic diameter was increased, which suggests systolic dysfunction (**Supp. Figure 1A-C**). Since reduced contractility was previously shown in animals with reduced mitochondrial gene expression, we hypothesized *CHCHD3/6* KD may reduce contractility via a role in mitochondrial function (Bhandari, Song and Dorn, 2015; Martínez-Morentin *et al*., 2015; Tocchi *et al*., 2015).

To test how early the cardiac phenotype of *CHCHD3/6* KD manifests in adult stages, 1-week old *Hand^4.2^-Gal4* KD of *CHCHD3/6* flies were examined, using several independent RNAi lines for *CHCHD3/6*. These flies also had reduced fractional shortening, i.e., reduced contractility due to systolic dysfunction (**Figure 1D-F**). This phenotype was observed in cardiac assays of intact flies (see Methods) (**Supp. Figure 1D-F**), as well as in the semi-intact adult heart preparation that lacks neuronal inputs (SOHA; (M Fink *et al*., 2009)). To further validate a cardiac-specific role for *CHCHD3/6*, as opposed to non-autonomous effects from other tissues, we performed KD of *CHCHD3/6* using *Dot*-Gal4 (expressed in pericardial cells, PC, which also express Hand), *Mef2* (Myocyte enhancer factor 2)-Gal4 (a pan-muscle driver), or *elav*-Gal4 (a pan-neuronal driver) (see Methods). A large reduction in fractional shortening was only observed with the pan-muscle driver that includes cardiac muscle, but not with the PC or neuronal drivers, confirming a cardiomyocyte-autonomous effect (**Supp. Figure 1J-L**). Both CHCHD3/6^RNAiA^ and CHCHD3/6^RNAiB^ lines had the same predicted off-target gene, *Duox* but *Hand^4.2^-Gal4* driven KD of *Duox* had no effect on fractional shortening, confirming that the cardiac effects were due to *CHCHD3/6* KD (**Supp. Figure 1M**).

### Temporal requirements for *CHCHD3/6* in maintaining heart function

We next sought to understand if *CHCHD3/6* has different temporal requirements for its effects on heart structure and function, since hearts of operated HLHS patients often develop reduced ejection fraction and heart failure, including the 11H proband. First, to assess whether *CHCHD3/6* could have a role in early heart development, we mined embryonic heart-specific single-cell transcriptomic data (Vogler *et al*., 2021) and found that *CHCHD3/6* was expressed in *Drosophila* cardioblasts (CBs), along with other cardiogenic factors (*tinman*, *H15*, and *Hand*) (**Figure 2A**). Next, we analyzed *CHCHD3/6* mutant embryos for cardiac phenotypes. Late-stage 16-17 *CHCHD^D1^* / *CHCHD^DefA^* trans-heterozygous embryos were stained for Mef2 (early mesoderm/muscle-specific transcription factor) and Slit (secreted protein in the heart lumen) but did not exhibit overt cardiac specification defects (**Figure 2B**). We used the *tinD*-Gal4 driver (*tinman enhancer D* (Yin, Xu and Frasch, 1997; Xu *et al*., 1998) to test whether *CHCHD3/6* KD in the dorsal mesoderm (including cardiac mesoderm) during embryonic stages 10-12 affects establishment of adult heart function. We reared *tinD-Gal4*>CHCHD3/6^RNAiA^ flies at 29°C throughout life to achieve high KD efficiency but did not observe reduced fractional shortening or any other functional defects, relative to controls (**Figure 2C, D**). Thus, KD of *CHCHD3/6* in the embryonic cardiac mesoderm is not sufficient to impact later heart function.

**Figure 2:**
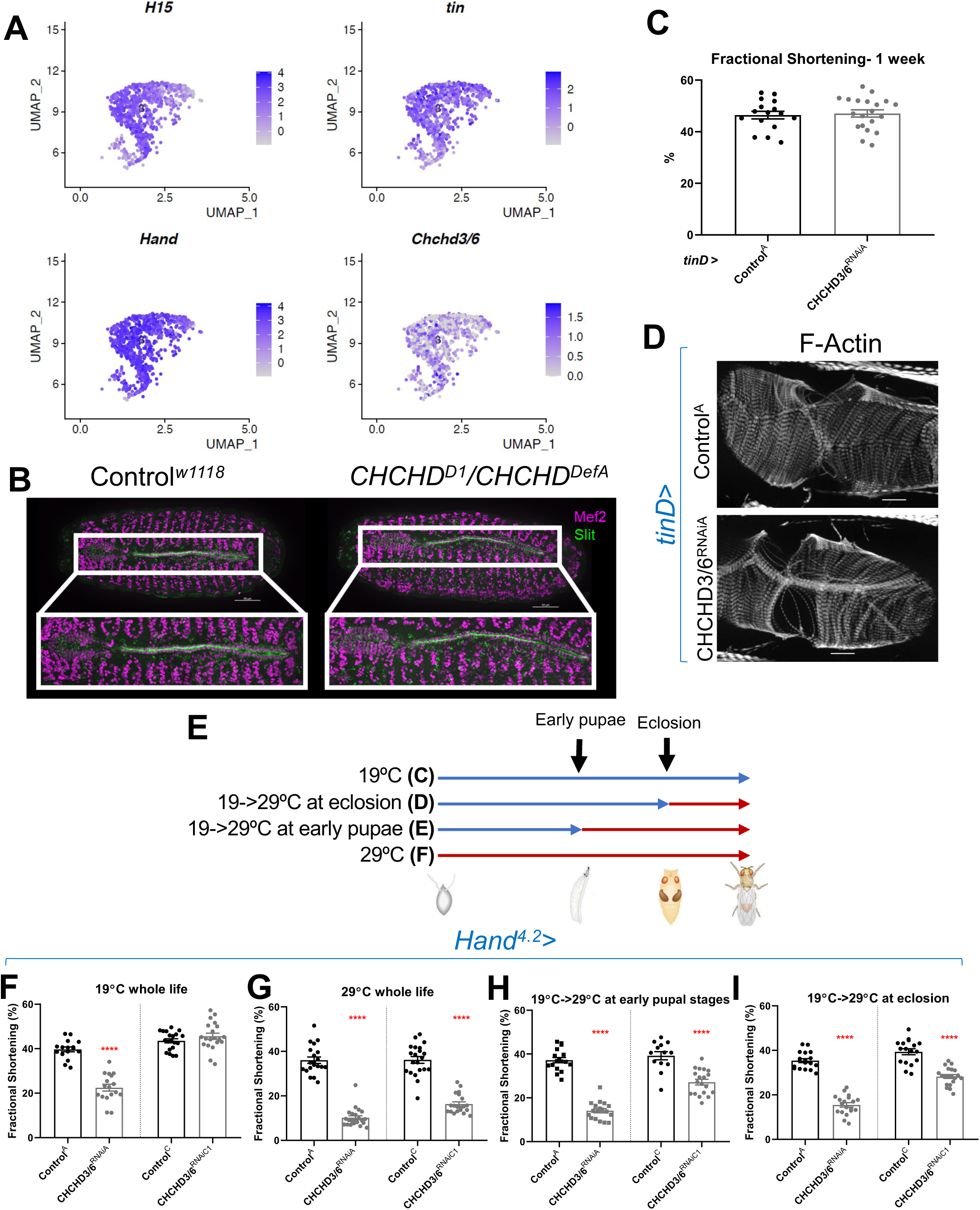
*CHCHD3/6* expression is important for adult cardiac function around larval stages and early adult stages. **A)** UMAP (uniform manifold approximation and projection) plot from CB-specific single-cell transcriptiomics (Vogler *et al*., 2021) showing expression of *CHCHD3/6* in CBs, as identified by cardiac TFs *tin*, *H15*, and *Hand*. **B)** Stage 16-17 embryos (late stage cardiogenesis) were collected from a *CHCHD3/6* loss of function line (*CHCHD^D1^*) line crossed to a *CHCHD3/6* deficiency line (*CHCHD^DefA^*) and stained for Mef2 (all muscle transcription factor, magenta) and Slit (secreted protein of the lumen, green). 50µm scale. **C)** *tinD*>Control^A^ or >CHCHD3/6^RNAiA^ were reared at 29°C and females were filmed and imaged at 1-week of age. A) *tinD*>CHCHD3/6^RNAiA^ did not have a significant reduction in fractional shortening compared to *tinD*>Control^A^ flies. **D)** F-actin was unchanged between *tinD*>Control^A^ and *tinD*>CHCHD3/6^RNAiA^ flies at 1 week of age; 20µm scale. **E)** Schematic overview of temperature shift experiments. **F-I)** Fractional shortening measurements from 1-week old female flies reared at **F)** 19°C for whole life, **G)** at 29°C for whole life, **H)** 19°C, and moved to 29°C at early pupal stages, or **I)** 19 °C, and moved to 29°C once eclosed (virgin flies), Unpaired two-tailed t-test, ****p≤ 0.0001, error bars represent SEM.

To further investigate the temporal requirement of *CHCHD3/6* during heart development, we made use of the temperature-dependence of Gal4-mediated KDs (less KD efficiency at 19°C, greater KD efficiency at 29°C; see **Figure 2E** for experimental strategy). *Hand^4.2^-Gal4* mediated KD of *CHCHD3/6*^RNAiA^ had strong contractility defects already at 19°C (**Figure 2F**). A weaker RNAi KD line *CHCHD3/6*^RNAiC1^ (see **Figure 1D-F**) caused no reduction in fractional shortening at 19°C (**Figure 2F**), whereas at 29°C fractional shortening was reduced similarly to the stronger KD line 19°C. To examine different developmental windows, *Hand^4.2^-Gal4*>*CHCHD3/6*^RNAiC1^ flies were shifted from 19°C to 29°C at either early pupal stages or early adult stages (after eclosion) until 1 week of age when heart function was assessed (**Figure 2E**). Interestingly, both treatments caused a substantial reduction in fractional shortening, although somewhat less than at 29°C throughout life (**Figure 2G-I**). This suggests that *CHCHD3/6* is not only required during pupal development, but also at adult stages for maintaining robust heart function.

### Cardiac knockdown of *Drosophila CHCHD3/6* results in severe reduction of sarcomeric Actin and Myosin levels

The strong heart functional defects upon *CHCHD3/6* KD suggest that the contractile machinery in cardiomyocytes is severely compromised. To probe for contractile abnormalities, we examined several sarcomeric components, including filamentous (F-) Actin, Myosin heavy chain, Obscurin (present at the M-line), α-Actinin (present at the Z-line), and Sallimus (Titin component in flies, localized near the Z-line) (**Figure 3A**). The intensity of F-actin staining with phalloidin was severely diminished in the working myocardium of *Hand^4.2^-Gal4*>*CHCHD3/6* KD flies (**Figure 3B-D**, arrows in **B**). Because *Hand^4.2^*-Gal4 expression is less in ostial cardiomyocytes (inflow valves), F-actin staining in ostial sarcomeres was minimally affected, if at all (**Figures 3B**, arrowheads). Like F-actin staining, Myosin staining was also dramatically diminished (**Figure 3E,I**) In contrast, Obscurin and α-Actinin staining was only moderately reduced (**Figure 3F,G,J,K**), and Sallimus staining was unaffected (**Figure 3H, L**). These findings suggest that loss of CHCHD3/6 function did not abrogate the overall sarcomeric organization, but instead differentially affected the abundance of individual sarcomeric proteins. Overall, the strongly diminished F-actin and Myosin levels in cardiac myofibrils is likely responsible for the diminished contractile capacity of the ATP-dependent actomyosin network in *CHCHD3/6* KD hearts.

**Figure 3:**
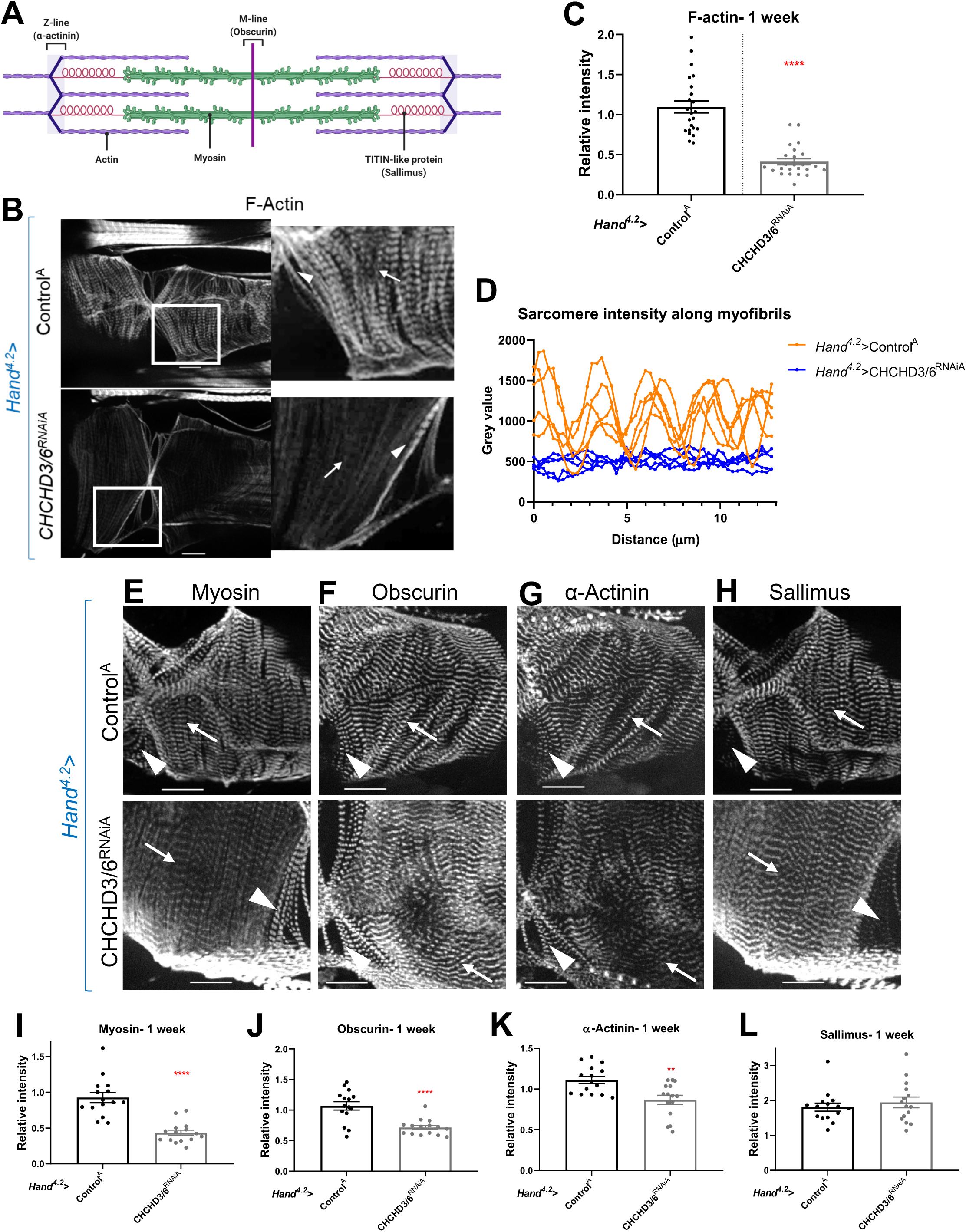
Cardiac tissue from heart-specific *CHCHD3/6* KD flies exhibit reduced and altered sarcomeric proteins in the myocardial tissue. **A)** Schematic of sarcomeric protein distribution inside myofibrils (image created with BioRender.com). **B)** F-actin staining in 1-week old female *Drosophila* hearts with *Hand^4.2^-Gal4* KD of *CHCHD3/6*. Arrowheads indicate ostial myofibrils and arrows point to myocardial myofibrils (non-ostial). **C)** F-actin intensity measured as mean gray value (gray value/# of pixels) along myocardial myofibrils relative to mean gray value of ostial myofibrils. **D)** Mean intensity of F-actin along individual myofibrils. 1-week old *Drosophila* hearts with *Hand^4.2^-Gal4* driven KD of control or *CHCHD3/6* stained for antibodies against **E)** Myosin, **F)** Obscurin, **G)** α-Actinin, or **H)** Sallimus. Arrowheads indicate ostial myofibrils and arrows point to working cardiomyocyte tissue (non-ostial). **I-L)** Mean fluorescence intensity along myocardial myofibrils relative to ostia myofibrils in 1-week old *Hand^4.2^-Gal4*>CHCHD3/6^RNAiA^ adults stained for sarcomeric proteins **I)** Myosin, **J)** Obscurin, **K)** α-Actinin, or **L)** Sallimus. Unpaired two-tailed t-test, **p≤0.01, ****p≤0.0001; error bars represent SEM. 20µm scale.

### Actin polymerization components do not mediate sarcomeric actomyosin reduction upon cardiac *CHCHD3/6* knockdown

Due to the strong reduction of F-actin levels observed with reduced *CHCHD3/6* expression, we hypothesized that globular (G) to F-actin polymerization was disrupted. If *CHCHD3/6* KD compromises mitochondrial ATP production in high energy-demanding CMs, the reduced ATP levels could disrupt actin polymerization and lead to reductions in F-actin and other sarcomeric proteins (Carlier, Pantaloni and Korn, 1984; Korn, Carlier and Pantaloni, 1987; Carlier, 1998). To test this, we reduced the cardiac expression of several actin polymerizing and depolymerizing genes (**Supp. Figure 2A)**. Cardiac KD of *Arp2/3*, *gel*, *Chd64*, *WASp*, and *TM1* caused slightly reduced fractional shortening, but not as severe as with *CHCHD3/6* KD (**Supp. Figure 2A, B**). Moreover, cardiac KD of *Arp*, *Chd64*, or *WASp* resulted in substantial myofibrillar disorganization, including gaps, but did not appear to produce the *CHCHD3/6* KD-like reduction in sarcomeric F-actin levels (WASp example shown in **Supp. Figure 2C**). Overall, KD of any of these genes involved in F-actin polymerization could not recapitulate the reduced myocardial F-actin intensity with normal sarcomeric patterning seen with *CHCHD3/6* KD. Therefore, it appears unlikely that defects in actin polymerization mediate the effects of *CHCHD3/6* KD.

### *CHCHD3/6* knockdown in flight or heart muscles results in defective mitochondria

Next, we hypothesized that the reduced contractile capacity and altered F-actin and Myosin in *CHCHD3/6* KD hearts was due to reduced mitochondrial function. We first examined mitochondrial integrity and sarcomeric actin staining in indirect flight muscles (IFMs), since their mitochondria are easily visualized due to their large size. Upon *CHCHD3/6* KD in IFMs using the pan-muscle driver *Mef2-*Gal4, we observed reduced F-actin staining and diminished sarcomere pattern definition (**Figure 4A**), similar to the cardiac phenotype (**Figure 3B**). This further indicated that the *CHCHD3/6* KD phenotype is not specific to cardiac tissue, but likely affects all muscles. We then examined mitochondrial integrity upon *CHCHD3/6* KD in IFMs expressing Mito::GFP (complex IV), and with antibodies against ATP synthase (complex V). Strikingly, Mito::GFP and ATP synthase staining revealed mitochondrial defects as manifested in aggregates (**Figure 4B-E**), which is suggestive of an imbalance between fusion and fission.

**Figure 4:**
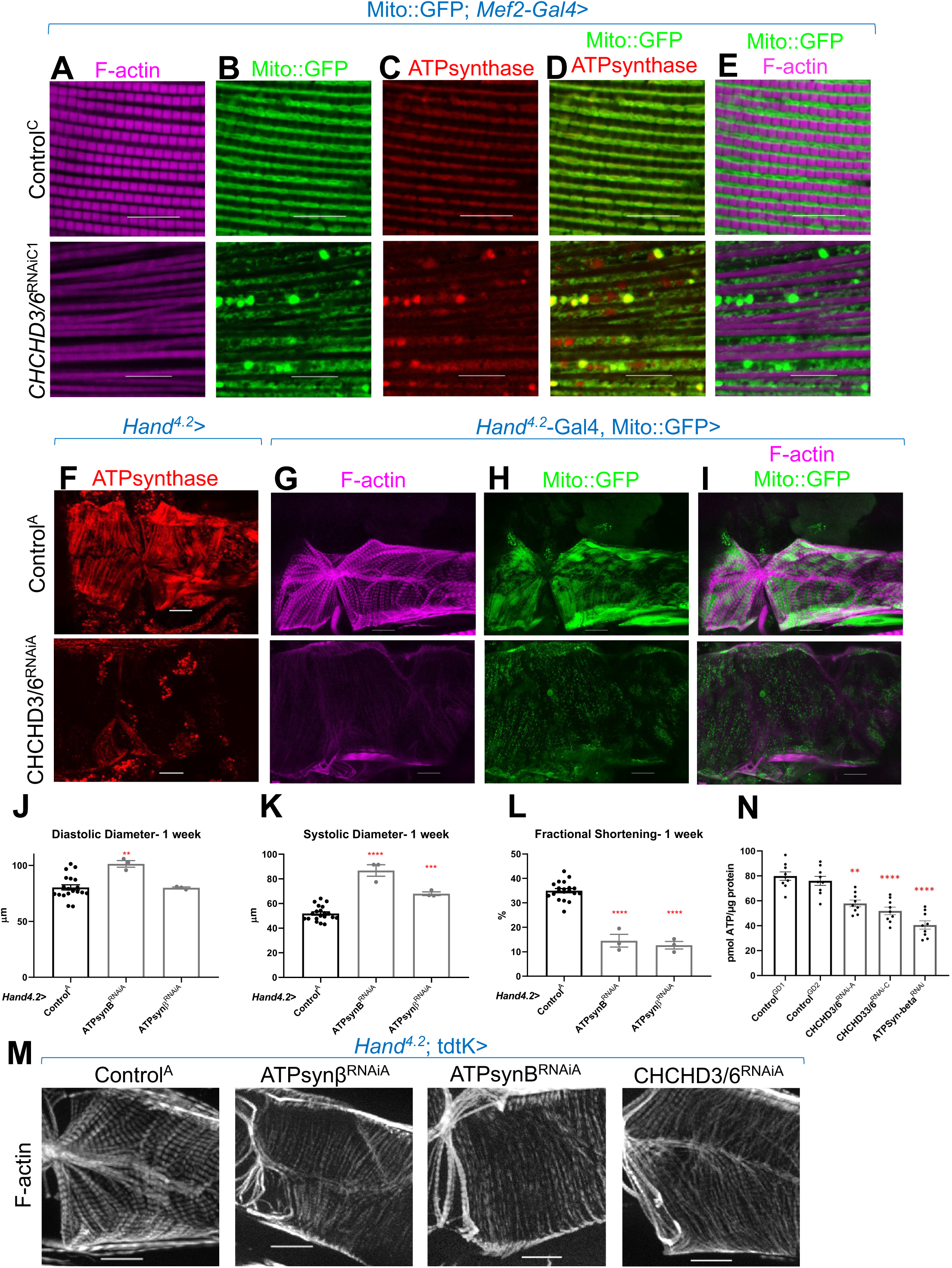
Mitochondria are aggregated and show reduced Complex IV and V staining upon loss of *CHCHD3/6*. Visualization of F-actin and mitochondria in *Drosophila* indirect flight muscles (IFMs). **A-E)** 1-2 day-old male *Drosophila* IFMs with Mito::GFP; *Mef2*-Gal4 stained for **A)** F-actin, **B)** GFP (Mito::GFP (GFP tagged COX8A)), **C)** ATP synthase, **D)** merged image of B + C, and **E)** merged image of A + B,10µm scale. **F)** *Hand^4.2^*>CHCHD3/6^RNAiA^ heart tissue at 1-week of age. **G-I**) F-actin and Mito::GFP staining in 1-week old female hearts using the *Hand^4.2^*-*Gal4*; Mito::GFP driver, 20µm scale. **J-L)** *Hand^4.2^-Gal4*; tdtK driven KD of ATP synthase subunits at 1-week of age measuring **J)** diastolic diameter, **K)** systolic diameter, and **L)** fractional shortening. Data is plotted as ± SEM and significance indicated relative to Control^GD2^. \*\*\*\**P ≤* 0.0001, \*\**P ≤* 0.01. **M)** 1-week old *Hand^4.2^-Gal4*; tdtK driven KD of ATP synthase subunits with altered F-actin (*CHCHD3/6* KD is depicted to contrast the structural phenotypes). **N)** Quantification of ATP levels from hearts of 1-week old flies (10-12 hearts per sample). ATP measurements were plotted relative to protein content. Statistical differences were calculated by one-way ANOVA followed by Tukey’s *post hoc* test for multiple comparisons.

Next, we assayed ATP synthase staining in *Hand^4.2^-Gal4*>*CHCHD3/6*^RNAiA^ hearts. We again observed mitochondrial aggregates, along with reduced F-actin and ATP synthase staining, relative to controls (**Figure 4F-I**). Furthermore, *Hand^4.2^-Gal4*, Mito::GFP*>*CHCHD3/6^RNAiA^ hearts also exhibited mitochondrial aggregates and reduced intensity of Mito::GFP (**Figure 4G-I**). Taken together, these data show that *CHCHD3/6* KD disrupts cardiac mitochondrial morphology and causes the formation of mitochondrial aggregates.

### Mitochondrial ATP synthase (complex V) KD causes similar contractile dysfunction and diminished sarcomeric F-Actin staining as with CHCHD3/6 KD

To determine whether KD of other mitochondrial genes impaired contractility and sarcomeric F-actin accumulation similar to that of *CHCHD3/6* KD, we screened RNAi lines from different mitochondrial functional groups (FlyBase.org GO term mitochondrion: 0005739) using the *Hand^4.2^-Gal-4*; tdtK driver for high-throughput heart imaging analysis (see Methods; Vogler, 2021). Cardiac KD of 17 of the 21 mitochondrial genes tested displayed reduced fractional shortening, most commonly due to systolic dysfunction (**Supplementary Table 2**). However, only KD of ATP synthase subunits reduced both fractional shortening and F-actin staining (**Figure 4J-M, Supp. Figure 3A**). Remarkably, F-actin staining in *Hand^4.2^-Gal4*, tdtK>ATPsynβ/B^RNAi^ hearts resembled that of *CHCHD3/6* KD, i.e. weakly stained myocardial myofibrils relative to ostial myofibrils (**Figure 4M**). These findings suggested that the heart dysfunction observed upon *CHCHD3/6* KD may be mediated via defects in ATP synthase.

### ATP production is reduced upon *CHCHD3/6* knockdown

Disrupted mitochondrial organization and reduced staining of OXPHOS components likely impacts ATP production and could explain cardiac functional and structural defects. We therefore directly measured ATP concentration in 1-week old female hearts with cardiac *CHCHD3/6* KD. We observed reduced ATP levels upon *CHCHD3/6* KD compared to controls, and these ATP levels were similar to those measured in response to cardiac *ATP-syn*βKD (**Figure 4N**). This further strengthens the hypothesis that the cardiac functional deficits in contractility induced by *CHCHD3/6* KD are due to mitochondrial defects that considerably reduce ATP levels in the heart necessary to build and maintain myofibrils.

### *CHCHD3/6* knockdown in all muscle cells is lethal or reduces climbing ability

To further characterize the impact of *CHCHD3/6* KD induced mitochondrial defects on muscle function, we assessed locomotive ability. When reared throughout development at 25°C, Mito::GFP; *Mef2*>CHCHD3/6^RNAiA^ flies were pupal lethal. However, with a moderate strength RNAi line (Mito::GFP; *Mef2*>CHCHD3/6^RNAiC1^) flies did eclose, but with reduced viability, especially for males (**Supp. Figure 3B**). Flies are negatively geotactic and will rapidly climb up the sides of a vial when tapped down. In 1-week of age, male and female Mito::GFP; *Mef2*>*CHCHD3/6*^RNAiC1^ flies, this activity was greatly reduced compared to controls (**Supp. Figure 3C**), supporting the hypothesis that *CHCHD3/6* KD reduces muscle function.

### Knockdown of SAM50 and Mitofilin cause cardiac defects, and SAM50 genetically interacts with CHCHD3/6

To further explore the role of the MICOS complex to maintain myofibrillar structure, we tested five MICOS complex-associated components for their requirement in cardiac contractility and sarcomeric F-actin levels (**Figure 5A**). *Hand^4.2^-Gal4*-mediated KD of *Mitofilin* or *Sam50* resulted in a significant reduction in fractional shortening, due to systolic dysfunction, mimicking *CHCHD3/6* KD defect in contractility, whereas *APOOL, Mic13* or *Mic10* KD did not exhibit significant effects (**Figure 5B, Supp. Figure 4A, B**). Of note, none of these five MICOS associated components displayed detectable reduction in sarcomeric F-actin staining upon KD (**Supp. Figure 4C**).

**Figure 5:**
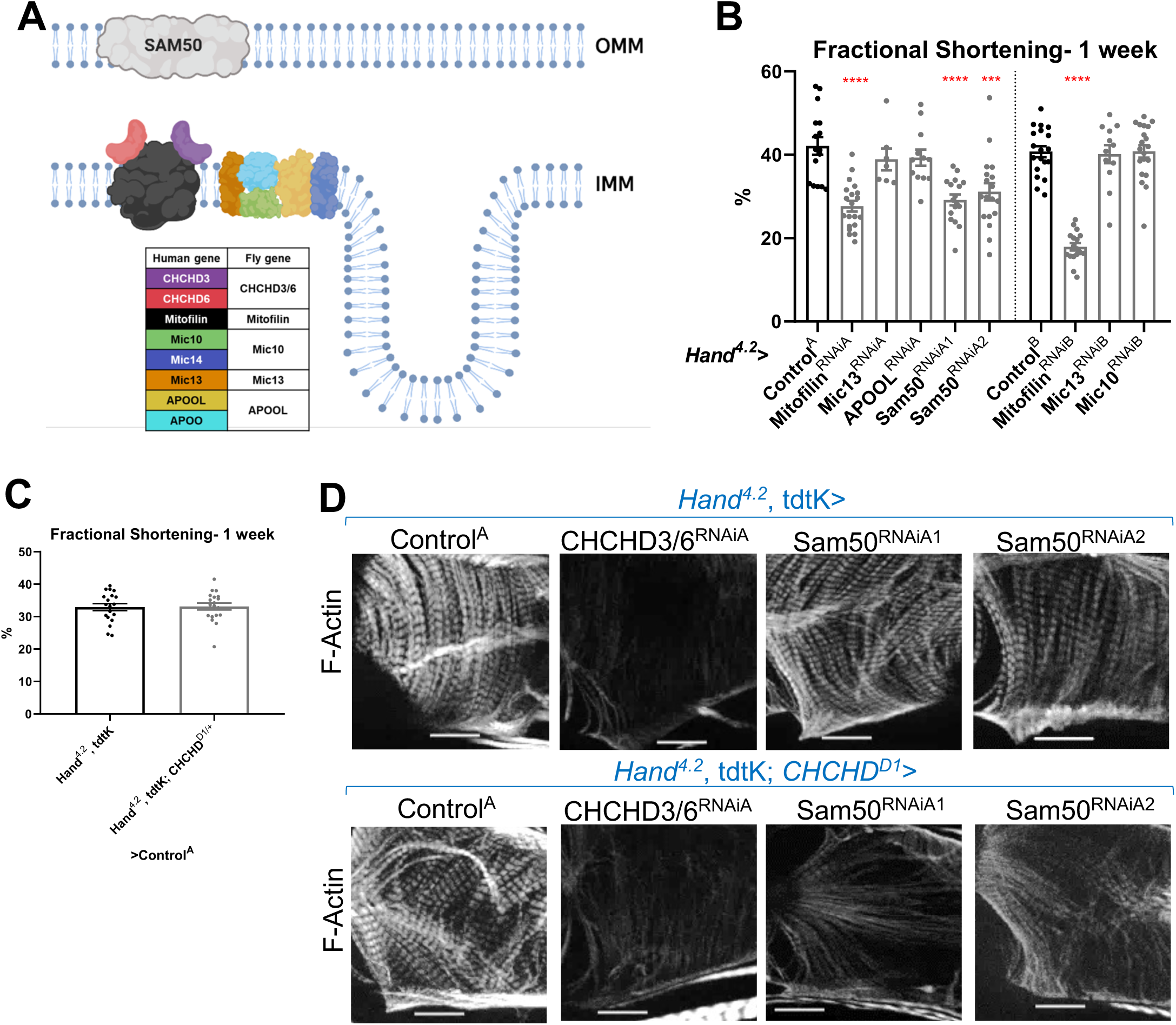
Assessment of other MICOS subunits in the *Drosophila* heart and in VCMs. **A)** Schematic of the MICOS complex and SAM50. Human MICOS subunits and their respective *Drosophila* homologs are listed (image created with BioRender.com). **B)** fractional shortening measured from 1-week old female flies with KD of individual MICOS subunits and *Sam50* using a *Hand^4.2^-Gal4* driver. Unpaired two-tailed t-test, ***p≤ 0.001, ****p≤ 0.0001; error bars represent SEM. **C)** *Hand^4.2^-Gal4*, tdtK; *CHCHD^D1/+^*line was crossed out with Control^A^. Unpaired two-tailed t-test, error bars represent SEM. **D)** 1-week old F-actin stained *Drosophila* hearts with or without heterozygous loss-of-function *CHCHD^D1/+^*in the background, 20µm scale.

Subsequently, we tested for genetic interactions between *CHCHD3/6* and MICOS-associated components, since protein-protein interactions among them have been previously described (Ding *et al*., 2015; Li *et al*., 2016; Tang *et al*., 2020). To test for interactions, we generated a *Hand^4.2^-Gal4, tdtK; CHCHD^D1/+^* heterozygote mutant sensitizer line that had no noticeable cardiac abnormalities (Deng *et al*., 2016) (**Figure 5C**). *Hand^4.2^-Gal4, tdtK; CHCHD^D1/+^* crossed to Mitofilin or *Sam50* RNAi did not further reduce fractional shortening beyond what was observed in response to KD of the individual genes (**Supp. Figure 4D**). However, we observed an interaction in the combined *Sam50* KD and *CHCHD^D1/+^*hearts, where F-actin levels were also strikingly reduced compared to the single KD, similar the *CHCHD3/6* KD (**Figure 5D**). This suggests that there is a threshold requirement of MICOS/Sam50, which when reached induces reduced contractility AND diminished F-Actin levels.

### Knockdown of MICOS subunits impairs proliferation of human iPSC-derived cardiomyocytes

We next tested the effects of KD of *CHCHD3/6* and other MICOS-associated components in human cardiomyocytes (CMs) derived from iPSC (hiPSC-CM). Since reduced CM proliferation is hypothesized to be a major contributing factor for the etiology of HLHS (Gaber *et al*., 2013; Liu *et al*., 2017; Theis *et al*., 2020), we focused on proliferation as our readout. We used small interfering RNA (siRNA) (as in (Theis *et al*., 2020)) to KD genes in hiPSC-CMs. We found that KD of *CHCHD6* and *CHCHD3*, as well as all other MICOS subunits and *SAM50*, significantly reduced their proliferation in an EdU incorporation assay (**Supp. Figure 5A, B**), supporting a potential link for *CHCHD6* and other MICOS subunits in HLHS pathogenesis.

### Testing of candidate genes prioritized in HLHS probands with *CHCHD3* or *CHCHD6* variants reveals novel genetic interactors

Since we found an essential role for *CHCHD3/6* in establishing heart structure and function in the *Drosophila* heart model with possible relevance for HLHS pathology, we assessed the presence of variants in additional HLHS family trios. Among the 183 Mayo Clinic HLHS family trios and pediatric cardiac genomics consortium (PCGC) databank (Jin *et al*., 2017), there were three probands with variants in *CHCHD6* (including 11H) and four with *CHCHD3* variants (**Supplementary Table 3**).

The relative abundance of rare, predicted damaging *CHCHD3/6* variants in the Mayo Clinic cohort, together with the postulated oligogenic nature of HLHS, led us to test for genetic interactions between CHCHD3/6 and other HLHS candidate genes. Specifically, we prioritized candidate genes with rare coding and regulatory variants identified in HLHS probands who also carried *CHCHD3-* or *CHCHD6*-variants (**Supplementary Table 4)**. We generated a *CHCHD3/6* sensitizer line, *Hand^4.2^-Gal4, tdtK; CHCHD3/6^RNAiC1^*, which at 21°C does not exhibit significant contractility deficits (**Figure 6A**). We screened 120 RNAi lines representing 60 candidate HLHS genes for genetic interactions and identified three hits that had contractility defects only when co-knocked down with *CHCHD3/6^RNAiC1^* at 21°C (**Figure 6B**). *Cdk12* (human ortholog: *CDK12*) has been shown to activate RNA polymerase II to regulate transcription elongation (Bartkowiak *et al*., 2010), *Goliath* (human ortholog: *RNF149*) encodes an E3 ubiquitin ligase that localizes to endosomes (Yamazaki *et al*., 2013) and *β-Spectrin* (human ortholog: *SPTBN1*) is a scaffolding protein that links the actin cytoskeleton to the plasma membrane. KD of *Cdk12* in combination with *CHCHD3/6* KD also led to greater lethality of elcosed flies at 1 week-of-age compared to individual gene KD (**Figure 6C**). While interestingly, we found that co-KD of *β-Spectrin* and CHCHD3/6^RNAiC1^ at 21°C was the only combination that also diminished F-actin and Myosin staining, similar to co-CHCHD3/6^RNAiA^ KD (**Figure 6D**). In summary, our approach using a sensitized screening strategy to interrogate genetic interactions between patient-specific candidate genes is a powerful tool to identify novel interactions that are potentially involved in the oligogenic ethology of HLHS and other CHDs.

**Figure 6:**
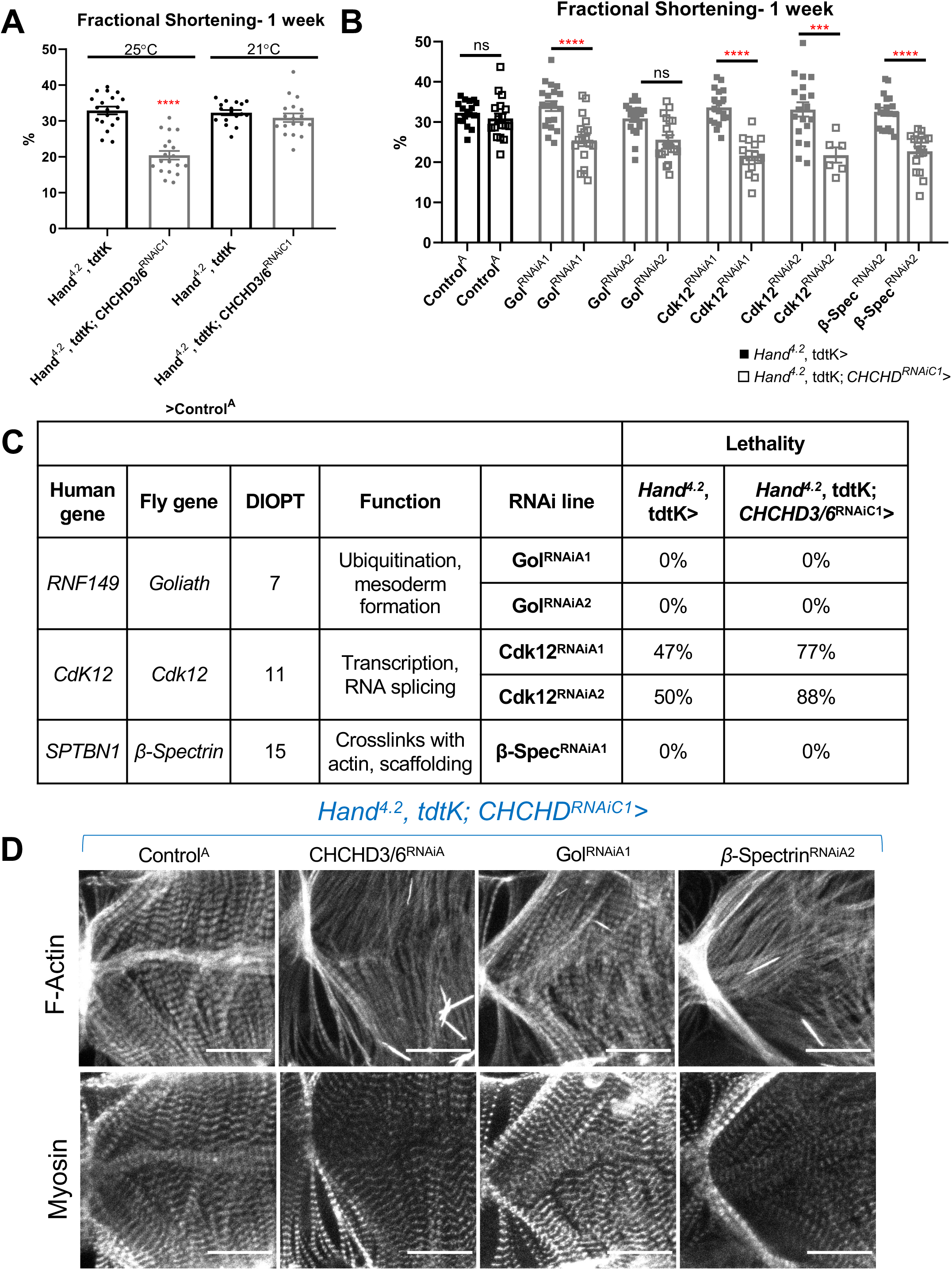
HLHS *CHCHD3* and *CHCHD6* family-based gene interaction screen reveals three hits. **A)** A *Hand^4.2^-Gal4*, tdtK; CHCHD3/6^RNAiC1^ sensitizer line show reduced fractional shortening at 25°C, which is no longer significant at 21°C. Unpaired two-tailed t-test, ****p≤ 0.0001; error bars represent SEM. **B)** Genetic interaction of CHCHD3/6 and prioritized HLHS candidates. Two-way ANOVA with Tukey’s multiple comparisons test, only statistical comparisons between the same RNAi lines are shown; ***p≤ 0.001, ****p≤ 0.0001; error bars represent SEM. **C)** Functional overview of human and *Drosophila* orthologs. KD of *Ckd12* with *CHCHD3/6* KD led to increased lethality of eclosed flies by 1 week-of-age.

## DISCUSSION

HLHS is characterized by a small left heart, including reduced left ventricle size and mitral and/or atrial atresia or stenosis, and aortic hypoplasia, collectively obstructing systemic blood flow (Tchervenkov *et al*., 2006). As a consequence, newborns cannot sustain systemic blood flow for more than a few days and therefore require treatment soon after birth. There is a need for improved therapies to treat HLHS patients, and this requires a better understanding of the biology behind HLHS pathogenesis. Here, we probed the genetic basis of HLHS using WGS and powerful bioinformatic gene variant prioritization in a large cohort of HLHS proband-parent trios combined with model system validation.

The 11H family was prioritized because of consanguinity, implicating a homozygous recessive mode of inheritance that resulted in a short list of nine candidate genes. These candidate genes were probed in *Drosophila* and iPSC-CMs for a potential role in cardiomyocyte development and function, to gain new insights into HLHS and CHDs in general. Among these HLHS gene candidates, we focused on *CHCHD3/6*, which has not been previously studied in the heart, and which had striking cardiac functional and structural defects in *Drosophila*. Specifically, the preliminary gene screen demonstrated that *CHCHD3/6* cardiac-specific KD caused reduced contractility and decreased sarcomeric F-Actin and Myosin staining.

Our data suggest that *CHCHD3/6* is necessary during larval and early adult stages to maintain contractility in the adult heart. This is relevant since patients with HLHS have both structural heart disease and risk for later myocardial failure (Theis, Zimmermann, *et al*., 2015). The prevailing“ no flow, no grow” hypothesis for HLHS pathogenesis surmises that reduced blood flow in the fetal heart causes underdevelopment of the left ventricle (Goldberg and Rychik, 2016; Grossfeld *et al*., 2019). A reduced ability for the heart to contract in utero, due to reduced *CHCHD6* activity, could contribute to decreased ventricular blood flow in the embryo, resulting in an abnormally small left ventricle. Moreover, reduced *CHCHD6* activity could compromise right ventricular function later in life (Theis, Hrstka, *et al*., 2015; Theis, Zimmermann, *et al*., 2015). In fact, the 11H proband exhibited mildly reduced right ventricular ejection fraction several years after successful surgical palliation. Consistent with our model system, *CHCHD6* deficiency could result in cumulative impairment of mitochondrial function, leading to contractile dysfunction (Sun, Youle and Finkel, 2016).

*CHCHD3/6* KD in the fly heart led to mitochondrial aggregates, with reduced ATP synthase (complex V) levels, and consequently impaired ATP production. Mitochondrial aggregates or fragments could be indicative of an imbalance between fission and fusion. It has previously been reported that *CHCHD3* KD in HeLa cells resulted in fragmented mitochondria that was due to improper mitochondrial fusion (Darshi *et al*., 2011). It has also been demonstrated in yeast that individual or combinatorial loss of MICOS complex proteins disrupt cristae morphology (Friedman *et al*., 2015), thus suggesting a mechanism by which *CHCHD3/6* loss could mediate HLHS pathogenesis. Furthermore, we identified a genetic interaction between SAM50 and CHCHD3/6 that leads to a contractile deficit and diminished sarcomeric F-Actin. Recent findings demonstrate that SAM50 directly interacts with Mic19, the mammalian ortholog of CHCHD3, to mediate inner and outer membrane bridging and cristae morphology (Tang *et al*., 2020).

Our data further suggest that ETC Complex V/ ATP synthase is a potential downstream effector of *CHCHD3/6* and MICOS complex function. Individual KD of ATP synthase subunits resulted in reduced fractional shortening and reduced sarcomeric actin. As a result, we hypothesize reduced *CHCHD3/6* expression affects ETC function, specifically ATP synthase, leading to reduced ATP production. OXPHOS complex assembly has been shown to be disrupted upon MICOS depletion, and we speculate ATP synthase may be disrupted when *CHCHD3/6* is reduced (Cogliati, Enriquez and Scorrano, 2016). Consistent with this, we observe depletion of ATP synthase levels upon *CHCHD3/6* KD.

Finally, we tested a potential oligogenic basis of HLHS in our family-based *CHCHD3* and *CHCHD6* interaction screen and identified three hits that reduced fractional shortening only in conjunction with *CHCHD3/6*, but not on their own. Co-KD of *Cdk12* and *CHCHD3/6* also reduced fractional shortening, and caused greater lethality relative to *Cdk12* KD alone. Cdk12 activates RNA polymerase II to regulate transcription elongation (Bartkowiak *et al*., 2010). We postulate that since *CHCHD3/6* is a nuclear-encoded gene, reducing transcription with *Cdk12* KD could decrease *CHCHD3/6* levels in a background where *CHCHD3/6* activity is already compromised. Alternatively, reduced transcription of other nuclear genes associated with ATP production in combination with *CHCHD3/6* KD could further reduce ATP levels enough to cause contractility defects. In support of this, a study examining the effects of RMP (RNA polymerase II subunit 5-mediating protein) found that mice with cardiac-specific *Rpm* KO exhibited reduced fractional shortening and ATP levels, which were attributed to a reduction in mRNA and protein levels of the mitochondrial biogenesis factor PGC1α (Zhang *et al*., 2019). The second hit, *Goliath*, is an endosomal ubiquitin E3 ligase. Although Goliath has been implicated in endosomal recycling (Yamazaki *et al*., 2013), its role in *Drosophila* mitophagy *in vivo* has not been examined. Reduced cardiac contractility with co-KD of *Gol* and CHCD3/6 could result from impaired mitophagy and reduced mitochondrial biogenesis. Together, the accumulation of damaged mitochondria can reduce ATP content required for contraction (Palikaras and Tavernarakis, 2014; Palikaras, Lionaki and Tavernarakis, 2015). The third hit, *β-Spectrin,* acts as a scaffolding protein. Recent data suggests that the human ortholog, SPTBN1 (Nonerythroid spectrin *β*) influences SPTAN1 (Nonerythroid spectrin α) levels, which has a calmodulin binding domain (Ackermann and Brieger, 2019). Therefore, decreased *β-Spec* expression could reduce Calmodulin levels, thereby reducing contractility due to the combined reduction in Ca^2+^ handling and *CHCHD3/6* KD-induced reduced ATP levels.

In summary, we have identified a novel mechanism potentially involved HLHS pathogenesis, starting by analyzing WGS data from a prioritized family and large cohort of HLHS patients, followed by functional testing *in vivo* using the *Drosophila* heart model and *in vitro* using human iPSC-derived CMs. Compromised contractile capacity, diminished sarcomeric F-Actin and Myosin accumilation, and mitochondrial dysfunction in *CHCHD3/6* KD *Drosophila* hearts are promising phenotypes that could contribute to early HLHS manifestations or heart failure complications later in life. Further examination of the interactions between the MICOS complex and other emerging candidate genes will identify novel gene functions and pathways that contribute to HLHS pathogenesis. Furthermore, a detailed elucidation of novel candidate genes and genetic interactions based on patient-specific rare potentially damaging variants is expected to lead to gene networks that are relevant for HLHS and other CHDs.

## Data Availability

All data produced in the present study are available upon reasonable request to the authors

## Acknowledgments

We gratefully acknowledge the patients and families who participated in this study. We thank Marco Tamayo and Bosco Trinh for excellent technical assistance. This work was supported by National Institutes of Health (R01 HL054732 to R.B.). This work was also supported by a grant from the Wanek Foundation at Mayo Clinic in Rochester, M.N., to J.L.T., T.J.N., T.M.O., R.B. and A.R.C.; by the American Heart Association: AHA Predoctoral Fellowship (18PRE33960593 to K.B.) and AHA Postdoctoral Fellowship (20POST35180048 to N.J.K.).

## Materials and Methods

### Study subjects

Written informed consent was obtained for the index family and HLHS cohort, under a research protocol approved by the Mayo Clinic Institutional Review Board. Cardiac anatomy was assessed by echocardiography.

### Comparative genomic hybridization

To detect aneuploidy, array comparative genomic hybridization was performed using a custom 180K oligonucleotide microarray (Agilent, Santa Clara, CA), with a genome-wide functional resolution of approximately 100 kilobases. Deletions larger than 200 kilobases and duplications larger than 500 kilobases were considered clinically relevant.

### Genomic and bioinformatics analysis of 11H family

Genomic DNA was isolated from peripheral white blood cells or saliva. WGS and variant call annotation were performed utilizing the Mayo Clinic Medical Genome Facility and Bioinformatics Core. For the family quintet, 101 base pair (bp) or 150 bp paired-end sequencing was carried out on Illumina’s HiSeq 2000 or HiSeq 4000 platforms, respectively. Reads were aligned to the hg38 reference genome using BWA version 0.7.10 (http://bio-bwa.sourceforge.net/bwa.shtml) and duplicate reads were marked using Picard (http://picard.sourceforge.net). Local realignment of INDELs and base quality score recalibration were then performed using the Genome Analysis Toolkit version 3.4-46 (GATK) (McKenna *et al*., 2010). SNVs and INDELs were called across all samples simultaneously using GATK’s Unified Genotype with variant quality score recalibration (VQSR) (Poplin *et al*., 2018).

Variant call format (VCF) files with SNV and INDEL calls from each family member were uploaded and analyzed using Ingenuity Variant Analysis software (QIAGEN, Redwood City, CA) where variants were functionally annotated and filtered by an iterative process. Annotated variants were subject to quality filters and required to pass Variant Quality Score Recalibration (VQSR) and have a genotype quality score ≥20. Variants were excluded if they were located in a simple repeat region identified using tandem repeats finder (Benson, 1999) or were found to have a minor allele frequency >1% in gnomAD v2.1 (Karczewski *et al*., 2020). Second, functional variants were selected, defined as those that impacted a protein sequence, canonical splice site, microRNA coding sequence/binding site, or transcription factor binding site within a promoter validated by ENCODE chromatin immunoprecipitation experiments (Raney *et al*., 2014). Third, using parental and sibling WGS data, rare, functional variants were then filtered for those that were homozygous recessive in the proband.

### Analysis of HLHS cohort for variants in the MICOS complex

WGS was performed on samples from 183 individuals with HLHS and 496 family members by the Mayo Clinic Medical Genome Facility or Discovery Life Sciences. SNVs and INDELs that passed quality control were subject to further filtering based upon rarity (MAF < 0.01) and predicted consequence. Details about the sequencing and subsequent variant filtering to identify HLHS gene candidates have been previously described (Theis *et al*., 2021). Rare variants from the 183 probands were interrogated for variants in CHCHD3 and CHCHD6 to identify variants that arose de novo or were homozygous recessive, compound heterozygous or X-linked recessive. Next, inherited variants in these genes were analyzed, but stricter thresholds were required to identify the most damaging variants. Missense variants were required to have CADD>24 (corresponds to the upper quartile of the most damaging missense variants) and non-coding variants were required to have a Position Weight Matrix (PWM) score >0.75 from the Factorbook database (selecting for variants predicted to disrupt canonical transcription factor binding sites). In addition to the Mayo Clinic HLHS cohort, the Pediatric Cardiac Genomics Consortium (PCGC) whole exome sequencing dataset was interrogated for candidate genes in the MICOS complex in patients with CHD (Jin *et al*., 2017).

### Analysis of *CHCHD3* and *CHCHD6* variant carriers

Using robust bioinformatics algorithms as previously described (Theis and Olson, 2022), a broad range of both family-based Mendelian inheritance modeling and cohort-wide enrichment analyses were applied to identify additional candidate genes in HLHS probands identified to have a rare, predicted-damaging coding or regulatory variant in *CHCHD3* or *CHCHD6*.

### *Drosophila* strains and husbandry

*Drosophila* crosses were reared and aged at 25°C, unless otherwise noted. *Drosophila* orthologs were determined using DIOPT (*Drosophila* RNAi Screening Center Integrative Ortholog Prediction Tool) which calculates the number of databases that predict orthology (out of a score of 16) (Hu *et al*., 2011). Fly stocks were obtained from Vienna Drosophila Resource Center (VDRC) and Bloomington Drosophila Stock Center (BDSC). Lines include *Hand^4.2^-Gal4* (Han *et al*., 2006), *tdtK* (Klassen *et al*., 2017), *tinCΔ4-Gal4* (Lo and Frasch, 2001), *Dot-Gal4* (Kimbrell *et al*., 2002), *Mef2-Gal4* (Ranganayakulu *et al*., 2010), Mito::GFP (BDSC: 8442), *CHCHD^DefA^* (BL: 26847)*, CHCHD3/6* A (VDRC: 52251), *CHCHD3/6* B (VDRC: 105329), *CHCHD3/6* C (BDSC: 51157), *CHCHD3/6* D (BDSC: 38984), DUOX (BDSC: 32903), Mitofilin A (VDRC: 47615), Mitofilin B (VDRC: 106757), Mic13 A (VDRC: 14283), Mic13 B (VDRC: 100911), Mic10 A (VDRC: 102479), APOO(L) A (VDRC: 31098), Sam50 A (VDRC: 33641), Sam50 B (VDRC: 33642) *CHCHD^D1^* was kindly shared by the Ge lab (Deng *et al*., 2016).

Assessment of lethality in co-KD of CHCHD3/6 and Cdk12 refers to percentage of surviving flies at 1 week-of-age, versus the number of flies eclosed on day 0.

### *In situ* heartbeat analysis

An *in-situ* dissection approach was used to expose the denervated beating fly heart (Martin Fink *et al*., 2009; Ocorr *et al*., 2009; Vogler and Ocorr, 2009). SOHA (Semi-automated optical heartbeat analysis) was used to analyze high speed video recordings to determine heart-related parameters (Martin Fink *et al*., 2009). Flies (n>15) were briefly anesthetized using filter paper with 10µm FlyNap and transferred to a 10 × 35mm Petri dish with Vaseline to attach the hydrophobic wing cuticle to the dish. Oxygenated room temperature artificial hemolymph (108mM NaCl, 5mM KCl, 2mM CaCl_2_•2H_2_O, 8mM MgCl_2_•6H_2_O, 15mM pH 7.1 HEPES, 1mM NaH_2_PO_4_•H_2_O, 4mM NaHCO_3_, 10mM sucrose, and 5mM trehalose) was added to each dish. Flies were dissected as per(Vogler and Ocorr, 2009) and oxygenated for minimum 15 minutes to equilibrate. Dissected flies were filmed for 30 seconds using an Olympus BX63 microscope (10X magnification), a Hamamatsu C11440 ORCA-flash4.0 OLT digital camera, and HCImageLive program. These videos were uploaded to SOHA (semi-automated optical heartbeat analysis), end diastolic and end systolic diameters were manually marked towards end of ostia, and heart-related parameters were extracted (Fink *et al*., 2009).

### *In vivo* heartbeat analysis

Norland #61 optical glue was placed on a 22 × 50mm coverslip (one small drop for each fly). Flies (n>15) were briefly anesthetized using filter paper with 10µm FlyNap, transferred to coverslip on individual adhesive drops with the dorsal side facing the coverglass, and cured for 30 seconds using ultraviolet light. The coverslip was then placed on a 10 × 35mm Petri dish and secured using putty. Fly hearts were filmed for 5 seconds using an Olympus BX63 microscope (20X magnification), a Hamamatsu C11440 ORCA-flash4.0LT digital camera, and HCImage Live program. All analysis was automatically processed using R (Vogler, 2021)(gvogler/FlyHearts-tdtK-Rscripts: First release of the R tdtK script. (Zenodo, 2021). doi:10.5281/zenodo.4749935.).

### Adult *Drosophila* heart immunohistochemistry

Flies were dissected as per (Vogler and Ocorr, 2009) in a 10 × 35mm Petri dish and EGTA was added to a final concentration of 10mM. EGTA was removed and replaced with 4% methanol-free formaldehyde for 20 minutes. Formaldehyde was removed and replaced with 1X PBS 3 times. Flies thoraxes were removed, abdominal walls were trimmed, and excess fat around heart was removed. Hearts were then washed 3 times with 0.3% PBTx (Triton-X) for 15 minutes on a shaker. PBTx was removed and replaced with 200µL 1° antibody solution (0.3% PBTx + 1° antibody), then a small piece of Parafilm with carefully placed over the solution to form and seal of liquid over the hearts. Dishes were incubated either 1) at 4°C overnight or 2) at room temperature for 2 hours while shaking. Once finished incubating, Parafilm was gently removed with forceps and 3 15-minute washes with 0.3% PBTx were performed. PBTx was removed and replaced with 200µL 2° antibody solution (0.3% PBTx + 2° antibody), then a small piece of Parafilm was carefully placed over the solution. Dishes were incubated either 1) at 4°C overnight or 2) at room temperature for 2 hours while shaking. Parafilm was gently removed with forceps and 3 15-minute washes with 0.3% PBTx were performed. PBTx was removed and replaced with 1X PBS. Ventral cuticle with attached hearts were carefully removed individually from the Vaseline layer and transferred to a 25 × 75 × 1mm slide with 2 18 × 18mm No. 1 coverslips glued to form a bridge and ProLong Gold antifade mounting medium (Invitrogen) in the middle. Flies were placed ventral side up and covered with a 18 × 18mm No. 1.5 coverslip, sealed with clear nail polish around the edges, and stored at room temperature for 24 hours until being moved to 4°C.

Primary Antibodies: anti-dMef2 (1:20, gift from Dr. Bruce Paterson); anti-slit (1:40, c555 DSHB); anti-Myosin (1:50, 3E8-3D3 DSHB); anti-Sallimus (1:100, Abcam); anti-ATP5A (1:100, Abcam 14748); anti-ATP5A1 (1:200, Invitrogen 43-9800). All Secondaries from Jackson Immuno Research Labs used at 1:500: Goat anti-Rat 594; Goat anti-Rabbit 647; Goat anti-Rabbit Cy5; Goat anti-mouse Cy3. Dyes: Phallodin 594 or 647 (1:100, Invitrogen).

### *Drosophila* indirect flight muscle dissection and immunofluorescence

Thoraxes were removed under light CO_2_ pressure and fixed for 40 minutes in 5% PFA, followed by three 2-minute PBS washes. IFM muscle fibers were removed using fine (#55) forceps and washed with 0.5% PBTx for 15 minutes, then washed with 0.1% PBTx twice for 15 minutes. All subsequent antibody stainings were diluted in 0.1% PBTx and incubated shaking at 4°C overnight to penetrate the muscle tissue. IFMs were transferred to a 25 × 75 × 1mm slide without a bridge. ProLong Gold antifade mounting medium (Invitrogen) was added, the samples were covered with a 18 × 18mm No. 1.5 coverslip, and sealed with clear nail polish around the edges.

### *Drosophila* embryo collection, fixation, and immunofluorescence

Adult flies were reared in a plastic bottle cage with a Petri dish on the bottom containing grape agar (Agar, EtOH, glacial acetic acid, grape juice) and yeast paste (yeast and H_2_O) at 25°C. After incubating (16 hours), embryos were carefully collected with a brush and placed in a mesh basket. Flies were washed with water, then placed in bleach for 3 minutes, followed by 30 seconds of wash with water. Embryos were removed from mesh and placed in a fixation solution (2 Heptane: 1 2X PBS: 1 10% formaldehyde) for 25 minutes. Formaldehyde layer (bottom) was removed, replaced with 500µL MetOH, vortexed, then the supernatant with vitelline membranes (middle layer) was removed, this was repeated once more. Embryos were washed with MetOH (3 rinses, followed by 1 hour on rotator). Embryos were stored in fresh MetOH at −20°C. 1° antibody was added and tube was placed on rotator at 4°C overnight. 1° was later removed using 3X15 minute 0.4% PBTx washes. PBTx was removed, replaced with 2° antibody (diluted in PBTx), and rotated for 2 hours. 2° was later removed using 3X15 minute 0.4% PBTx washes, then left in PBS at 4°C. Since *CHCHD^D1^* and *CHCHD^DefA^* are both homozygous lethal at adult stages, each line was rebalanced over TM6b YFP (*CHCHD^D1^*/TM6b YFP and *CHCHD^DefA^*/TM6b YFP). The YFP lines (*CHCHD^D1^*/TM6b YFP and *CHCHD^DefA^*/TM6b YFP) were crossed out and embryos were selected against GFP to obtain only *CHCHD^D1^*/*CHCHD^DefA^*embryos.

### Fixed sample imaging

Samples were imaged at 10X, 25X, or 40X magnification using a Zeiss Apotome.1 Imager Z1, a Hamamatsu C11440 ORCA-flash4.0 OLT digital camera, and Zeiss ZEN. In order to obtain higher resolution, confocal microscopy was performed for all immunohistochemistry experiments involving Mito::GFP and ATP synthase staining.

### Climbing assay

Flies were initially anesthetized using FlyNap, placed into 5 separate vials, and counted for a total at week 0 for Control C females= 178, Control C males= 125, CHCHD3/6 C females= 154, CHCHD3/6 C males= 112. Each week, flies were transferred using a funnel to a clean longer tube with no food and these tubes were placed in a Styrofoam cutout to hold the tubes for tapping. The vial holder was tapped down multiple times until flies were at the base of the vial and then left to record the percentage of flies which reached 10cm after 10 seconds. The vial holder was tapped down multiple time to achieve biological replicates= 4 and different batches of genotypes were examined for technical replicates= 5.

### Statistical analyses

All statistical analyses were performed using GraphPad Prism version 8.0.1 for Windows, GraphPad Software, San Diego, California USA, www.graphpad.com. Statistical tests used are stated in figure legends. T-tests were performed on most heart assays where only one variable was defined. For tdtK analyses, a ranked one-way ANOVA Kruskal–Wallis test was used. Combinatorial KD assays with MICOS subunits and *CHCHD3/6* loss-of-function in *Drosophila* were analyzed with two-way ANOVA.

### PCR

The *CHCHD^D1^* line was confirmed via PCR using primers CHCHDD1 F4: ATATATCCGACGATGTGG and CHCHDD1 R4: AGCTCCTGGTTCATCTGG (Q5® High-Fidelity 2X Master Mix New England Bio).

### Quantitative real time polymerase chain reaction (qRT-PCR)

RNA was extracted using Qiagen miRNeasy® Mini Kit and cDNA was synthesized with Qiagen QuantiTech® reverse transcription kit. qRT-PCR analysis was performed using Roche FastStart Essential DNA Probes Master and Roche LightCycler® 96 with 2 biological replicates and 3 technical replicates. Data was analyzed in the LightCycler® application. Primers include CHCHD3/6 F: GCTAGAGGAACTTCAAAGATGG, CHCHD3/6 R: GGGATAGGAGGATACTTTCGG, RP49 F: GCTAAGCTGTCGCACAAATG, RP49 R: GTTCGATCCGTAACCGATGT.

### Human iPSC-derived cardiomyocyte proliferation assays

At day 25 of differentiation, human iPSC-derived cardiomyocytes (hiPSC-CMs) were dissociated with TrypLE Select 10X (Gibco) for up to 12 min and action of TrypLE was neutralized with RPMI supplemented with 10% FBS. Cells were resuspended in RPMI with 2% KOSR (Gibco) and 2% B27 50X with vitamin A (Life Technologies) supplemented with 2 µM Thiazovivin and plated at a density of 5.000 cells per well in a Matrigel-coated 384-well plate. hiPSC-CMs were then transfected with siRNA (Dharmacon) directed against each gene using lipofectamine RNAi Max (ThermoFisher). Each siRNA was tested in quadruplicate. 48 hours post-transfection, cells were labeled with 10 µM EdU (ThermoFisher). After 24h of EdU incubation, cells were fixed with 4% paraformaldehyde for 30 minutes. EdU was detected according to the protocol and cells were stained with cardiac specific marker ACTN2 (Sigma A7811, dilution 1:800) and DAPI. Cells were imaged with ImageXpress Micro XLS microscope (Molecular Devices) and custom algorithms were used to quantify EdU+ hiPSC-CMs.

### ATP Measurements

Measurements of ATP were performed using a luciferase assay as described previously (Liu and Lu, 2010; Zanon *et al*., 2017). 10-12 hearts per sample were collected from 1-week old flies and homogenized in 100µl extraction buffer (100 mM Tris and 4 mM EDTA, pH 7.8) containing 6M guanidine-HCl followed by rapid freezing in liquid nitrogen. The samples were boiled for 5min and cleared by centrifugation at 14,000 x g. Supernatants were diluted 1:50 and ATP levels were determined using ENLITEN® ATP Assay System (Promega, Cat #FF2000) as per manufacturer instructions. Total protein levels were determined by BCA method (Pierce, Cat #23225). ATP measurements were normalized to protein.

### Hybridization Chain Reaction (HCR)

Hearts were exposed as described above and RNA in situ performed and analyzed as described in (Kirkland *et al*., 2021). Briefly, hearts were relaxed using 10mM EGTA in artificial hemolymph and fixed in 4% formaldehyde in 0.1% Tween 20-PBS for 20 minutes. Hearts were then washed with 0.1% Tween 20, PBS, 2 x 5 minutes. On ice, hearts were incubated in a methanol gradient with PBS for 5 minutes each (25%, 50%, 75%, 100%, 75%, 50%, 25%). Hearts were then permeabilized in 1% Triton 100-X in PBS for 2 hours at room temperature. The hearts were post-fixed with 4% formaldehyde in 0.1% Tween 20-PBS for 20 minutes at room temperature before washing on ice with 0.1% Tween 20-PBS, 2 x 5 minutes. Subsequently, samples were washed with 50% 0.1% Tween 20-PBS and 50% 5XSSCT (5X SSC, 0.1% Tween 20, H_2_O) for 5 minutes on ice, followed by 5X SSCT for another 5 minutes. The hearts were then transferred to a 96 well plate and the hearts incubated in probe hybridization buffer (Molecular Instruments) for 5 minutes on ice, then 30 minutes at 37°C. The solution was then replaced with 2µl of each probe in 200µl of probe hybridization buffer and incubated at 37°C overnight (up to 16 hours). Next, 4 x 15 minute washes were performed with probe wash buffer (Molecular Instruments) at 37° C, then 2 x 5 minute 5XSSCT and 1 x 5 minute amplification buffer (Molecular Instruments). 2µl of corresponding h1 and h2 hairpins (Molecular Instruments) were heated to 95°C for 90 seconds, cooled in the dark for 30 minutes and added to 100µl of amplification buffer. Hairpin solution was then incubated with the heart samples at room temperature overnight (up to 16 hours), in the dark. Next, samples were washed 2 x 5 minutes with 5X SSCT; 2 x 30 minutes with 5X SSCT; 1 x 5 minutes with 5X SSCT and rinsed 3 x with PBS. DAPI in PBS (1:500) was incubated with the samples for 15 minutes, then samples were again rinsed 3 x 5 minutes in PBS. Samples were then mounted and imaged as described above. To quantify expression, a maximum projection image was created in ImageJ from the confocal stack image and binarized. The region around the cardiomyocyte nucleus was traced and the ROI copied to the binary image for particle analysis. Since segmentation was imperfect for transcripts very close together and to account for differences in pocket size, the % area covered by the transcripts was used to assess statistical significance in Prism (Graphpad).

---

**Supp. Figure 1:**
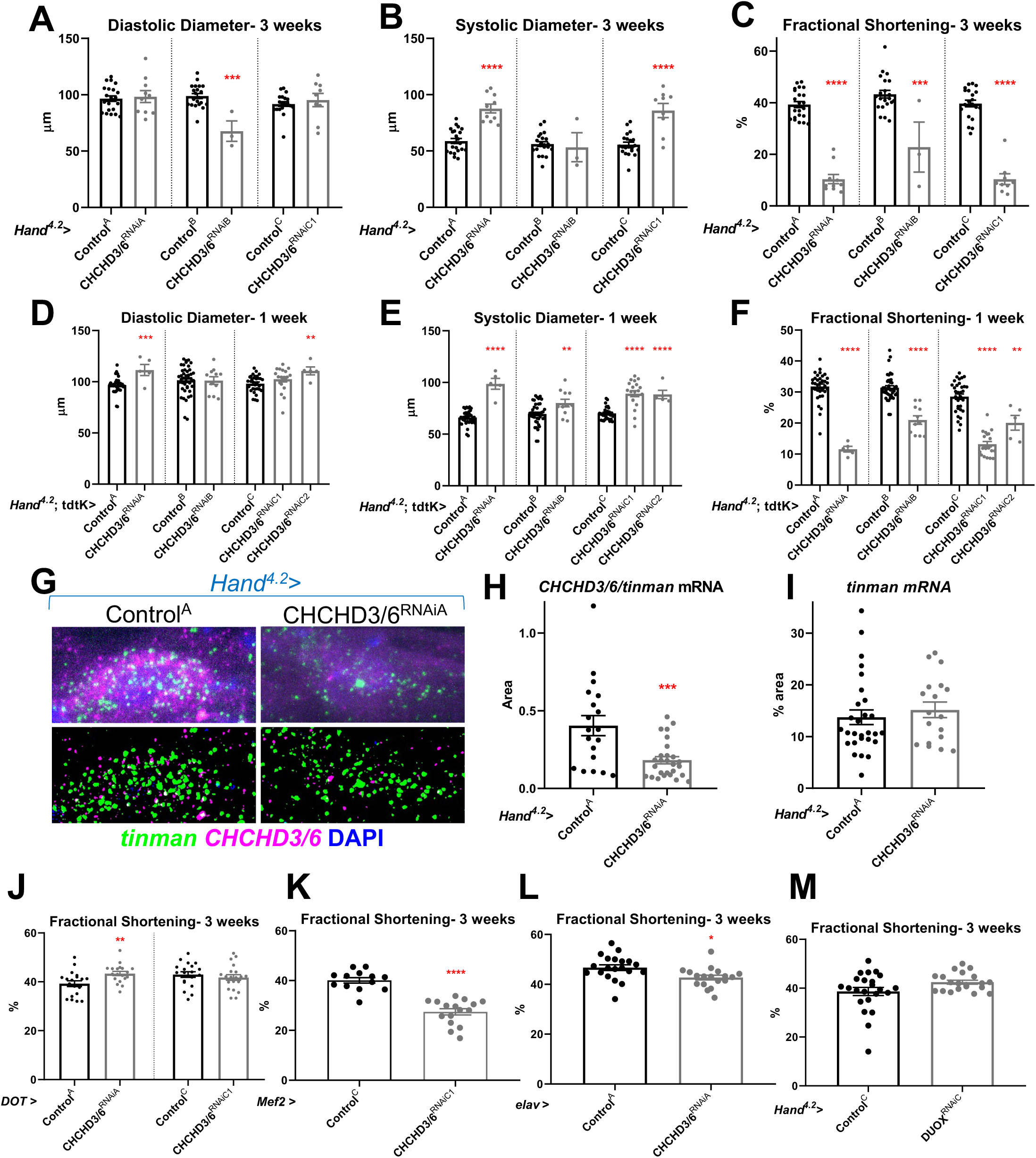
*CHCHD3/6* KD in the *Drosophila* heart causes reduced fractional shortening due to systolic dysfunction. **A)** EDD, **B)** ESD and **C)** FS from 3-week old *Hand^4.2^*>*CHCHD3/6* female flies. Very low number of *Hand^4.2^*>CHCHD3/6^RNAiB^ flies reaching 3 weeks-of-age indicates lethality possibly due to stronger KD of *CHCHD3/6*. **D-F)** 1-week old *Hand^4.2^-Gal4*, tdtK>*CHCHD3/6* female flies measured for **D)** EDD, **E)** ESD and **F)** FS. **G)** Confocal images of *CHCHD3/6* mRNA and *tinman* mRNA (top) and processed images (bottom) using a custom ImageJ macro. **H-I)** *CHCHD3/6* and *tinman* mRNA in 1-week old female *Drosophila* hearts. **H)** *CHCHD3/6* mRNA relative to *tinman* mRNA is reduced in *Hand^4.2^*>CHCHD3/6^RNAiA^ heart tissue. **I)** *tinman* mRNA % area, used as marker to normalize expression to. Fractional shortening in control and *CHCHD3/6* KD female flies at 3 weeks of age with a **J)** pericardial cell-specific driver (*DOT*-Gal4), **K)** all muscle cell specific driver (*Mef2*-Gal4), and **L)** neuronal driver (*elav*-Gal4). Note that *Mef2*>*CHCHD3/*6^RNAiA^ was lethal at pupal stages. **M)** Fractional shortening measurements from *Hand^4.2^*>DUOX^RNAiC^ 3-week old flies. Unpaired two-tailed t-test, *p≤0.05, **p≤0.01, ***p≤ 0.001, ****p≤ 0.0001; error bars represent SEM.

**Supp. Figure 2:**
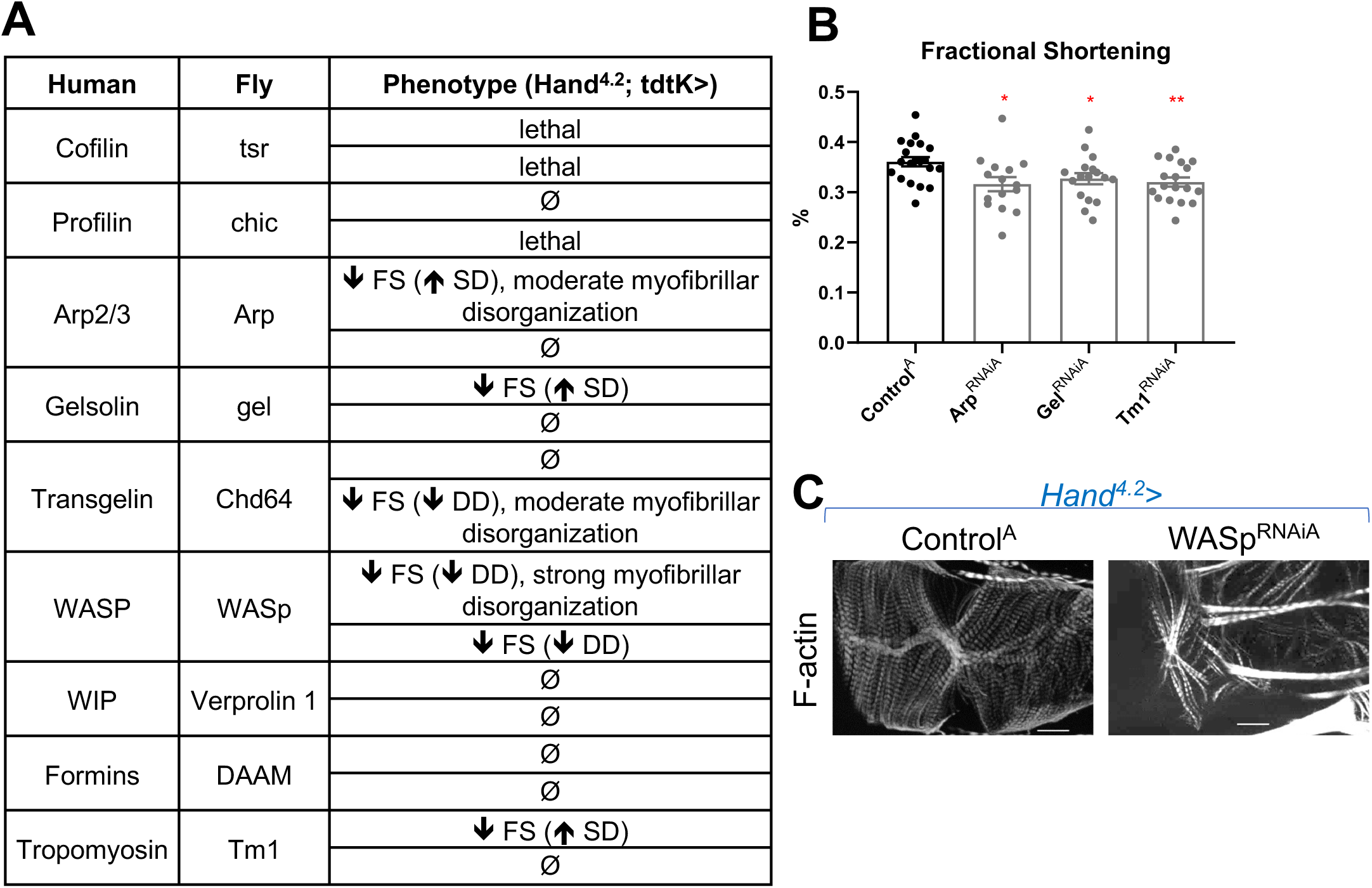
**A-C)** Candidate genes involved in polymerization/de-polymerization of F-actin phenotypes upon KD using a *Hand^4.2^*-Gal4;tdtK^attP2^ driver, measured at 1-week of age. **D)** Fractional shortening from RNAi^A^ lines. **E)** F-actin phenotype of hits **C)**. 20µm scale. Unpaired two-tailed t-test, *p≤0.05, **p≤0.01; error bars represent SEM. FS= fractional shortening, DD= End diastolic diameter, SD= Systolic diameter.

**Supp. Figure 3:**
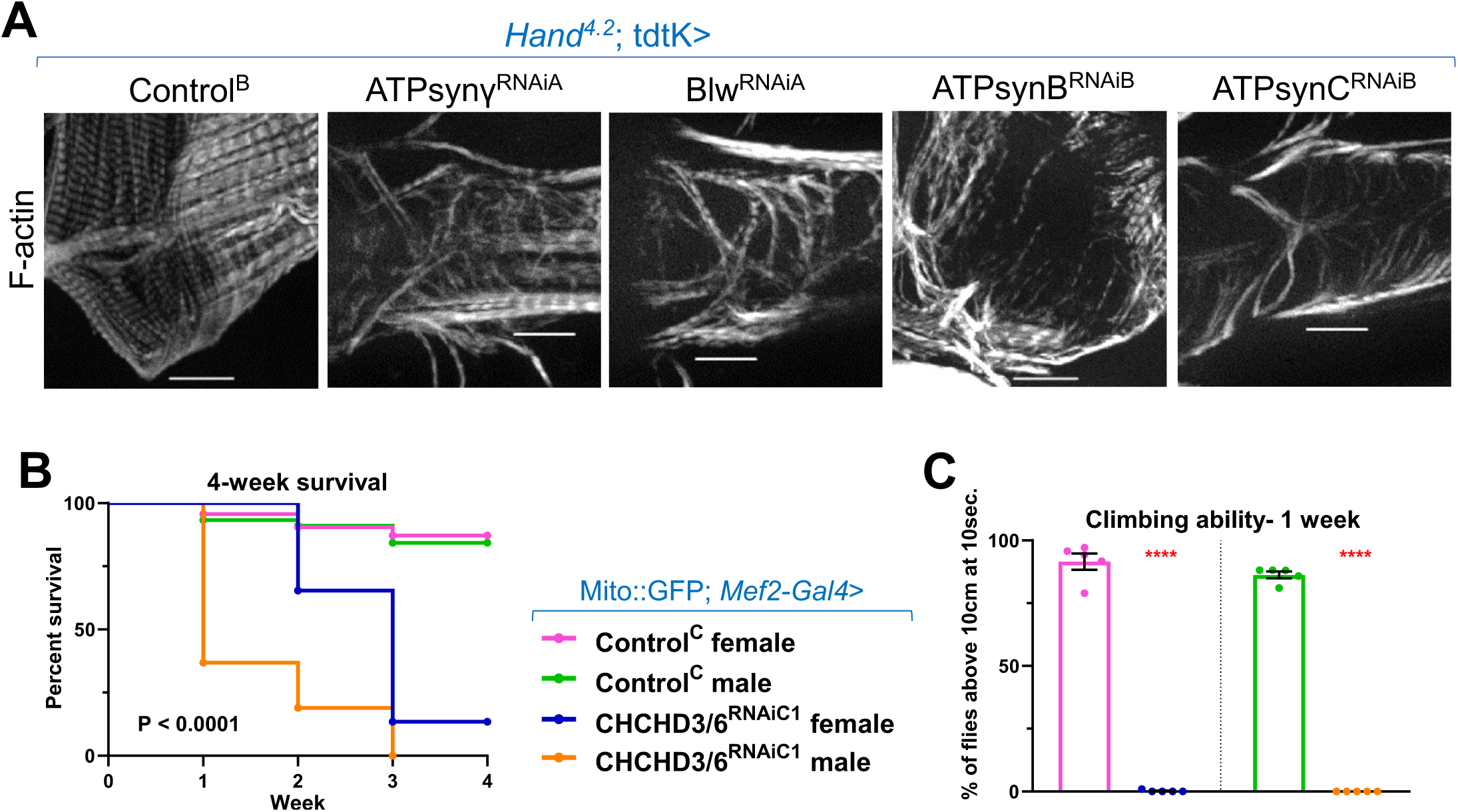
**A)** KD of mitochondrial ATP synthase subunits using *Hand^4.2^-Gal4*; tdtK produced strong F-actin phenotypes. 20µm scale. **B-C)** *CHCHD3/6* was knocked down using a Mito::GFP; *Mef2*-Gal4 line; note that Mito::GFP; *Mef2*-Gal4>CHCHD3/6^RNAiA^ is pupal lethal. **B)** Viability was significantly reduced in male and female CHCHD3/6^RNAiC1^ flies over a 4-week time course. Mantel-Cox test. **C)** Negative Geotaxis assay (average number of flies above a 10cm mark after being tapped down in a long vial) measured in 1-week old male and female flies. Unpaired two-tailed t-test, ****p≤ 0.0001; error bars represent SEM.

**Supp. Figure 4:**
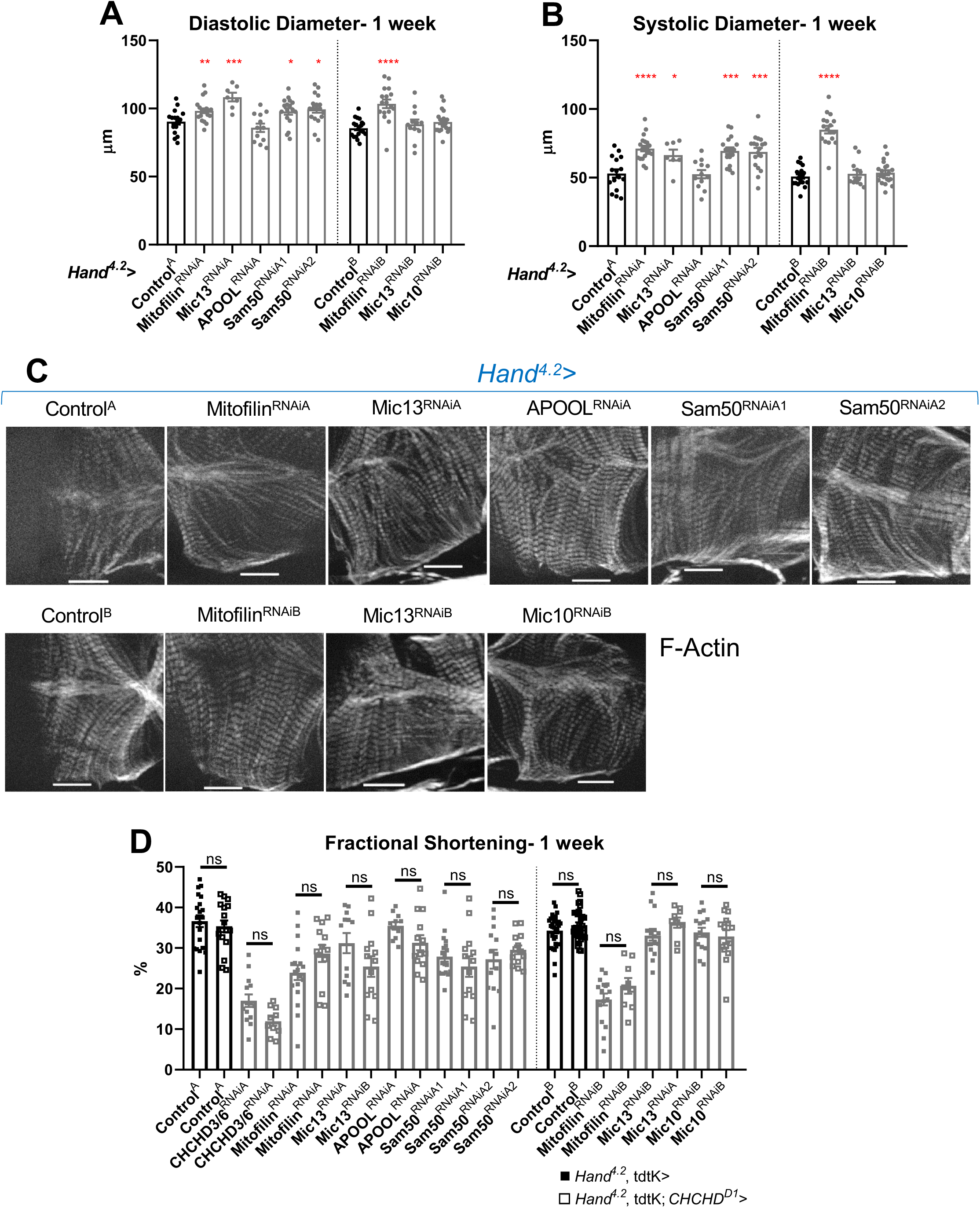
**A)** End-Diastolic diameter and **B)** End-systolic diameter measured from 1-week old female flies with KD of individual MICOS subunits and *Sam50* using a *Hand^4.2^- Gal4* driver. **C)** F-actin cardiac sarcomere staining in 1-week old female flies with *Hand^4.2^- Gal4* KD. **D)** All MICOS subunits and *Sam50* were knocked down with *Hand^4.2^-Gal4*, tdtK (filled squares) or using the sensitizer line *Hand^4.2^-Gal4*, tdtK; *CHCHD^D1/+^* (empty squares) at 25°C. Two-way ANOVA with Tukey’s multiple comparisons test, only statistical comparisons between same RNAi lines are shown; error bars represent SEM, 20µm scale.

**Supp. Figure 5:**
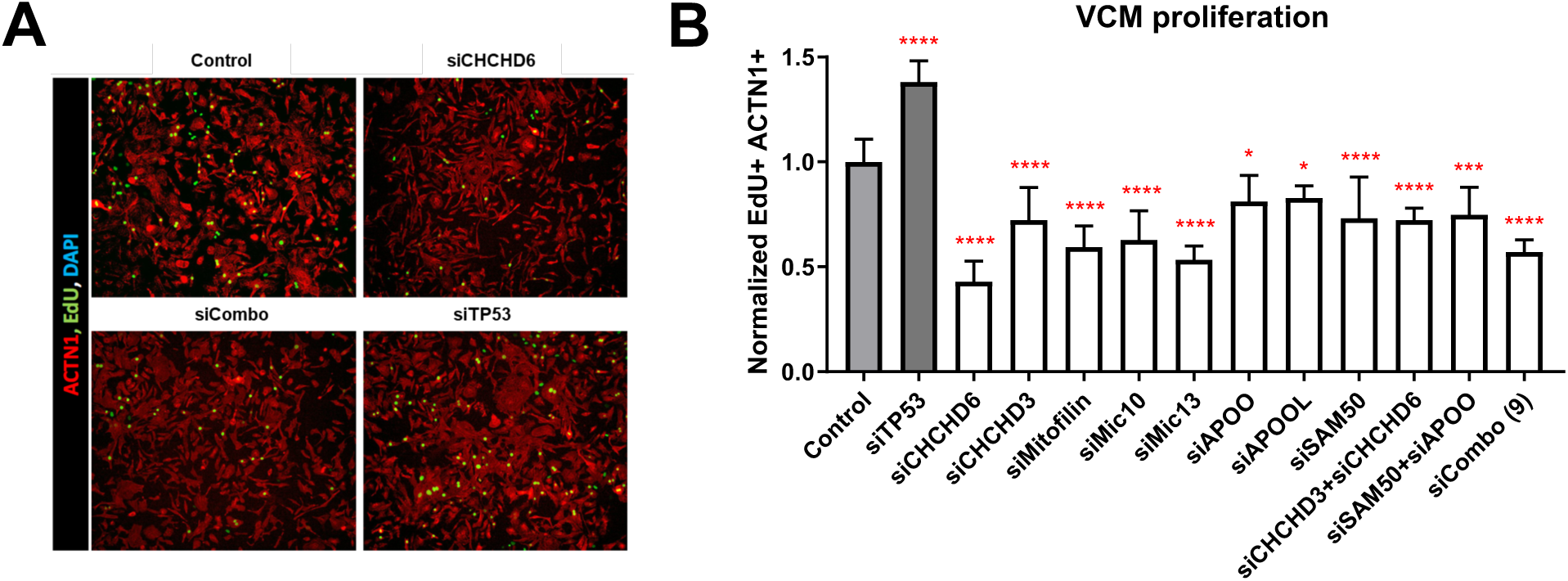
**A, B)** Cardiac cell proliferation assay following MICOS complex subunit KD, individually and in combination using EdU (proliferation marker, green) and ACTN1 (cardiac-specific, red) in ventricle-like cardiomyocytes (VCMs). **A)** VCMs showing reduced proliferation with si*CHCHD6*. **B)** Quantification of proliferation between siRNA mediated KD of individual MICOS subunits and MICOS subunit combination KDs in VCMs (siTP53 is a positive control). One-way ANOVA with Dunnett’s multiple comparisons test, *p≤0.05, ***p≤ 0.001, ****p≤ 0.0001; error bars represent standard deviation. One-way ANOVA with Dunnett’s multiple comparisons test, *p≤0.05, ***p≤ 0.001, ****p≤ 0.0001; error bars represent standard deviation.

**Supplementary Table 1:** Prioritized candidate genes from HLHS proband. Nine candidate genes harbored rare homozygous variants predicted to impact protein structure or gene regulation. Chr = chromosome, CADD = combined annotation dependent depletion, Hom = homozygous, Het = heterozygous, WT = wildtype, gnomAD = Genome Aggregation Database, MAF = minor allele frequency.

| Gene symbol | Chr | BP Position (HG38) | Transcript variant | Gene Region | Protein variant | rsID | RegulomeDB Rank for Regulatory Variants | CADD Score for Missense Variants (Percentile) | Genotype in sister | Genotype in brother | gnomAD MAF (%) |
| --- | --- | --- | --- | --- | --- | --- | --- | --- | --- | --- | --- |
| RHBDL2 | 1 | 38941780 | c.-224C>T | 5'UTR |  | rs570057342 | 4 |  | Hom | Het | 0.05 |
| CAP1 | 1 | 40039733 | c.-1061C>T; c.-1079C>T | Promoter |  | rs1033279388 | 4 |  | Hom | Het | not reported |
| SZT2 | 1 | 43427349 | c.3331G>T | Exonic | p.D1111Y | rs887336448 |  | 27 (85) | Hom | Het | not reported |
| RNF149 | 2 | 101308657 | c.-69C>G | 5'UTR |  | rs78958738 | 4 |  | Het | Hom | 0.65 |
| CHCHD6 | 3 | 126704425 | c.87+26C>A | Intronic |  | rs768047544 | 2b |  | Het | Hom | 0.00 |
| MTRR | 5 | 7889130 | c.1263C>G; c.1182C>G | Exonic | p.D394E; p.D421E | rs149678769 |  | 8 (15) | Het | Hom | 0.05 |
| MBTPS1 | 16 | 84056017 | c.2950G>A | Exonic | p.D984N | rs146299475 |  | 24 (65) | WT | WT | 0.66 |
| DGKE | 17 | 56833747 | c.-532A>C | Promoter |  | rs911884815 | 4 |  | WT | Het | 0.00 |
| C17orf67 | 17 | 56833747 | c.-1131T>G | 5'UTR |  | rs911884815 | 4 |  | WT | Het | 0.00 |

**Supplementary Table 2:** Mitochondrial gene screen in the *Drosophila* heart. RNAi lines of different mitochondrial functional groups were selected using the FlyBase.org gene ontology (GO) term mitochondrion (GO:0005739). The RNAi lines were crossed to *Hand^4.2^-Gal4*, tdtK and their progeny were assessed for contractility defects at 1-week of adult age using *Hand^4.2^-Gal4*; tdtK. Note that structural phenotype refers to any visible F-actin phenotype, not specifically to *CHCHD3/6* KD F-actin phenotype. FS= fractional shortening, DD= End diastolic diameter, SD= systolic diameter.

| Mitochondrial functional group | Gene | RNAi line | Functional phenotype | Structural (F-actin) phenotype |
| --- | --- | --- | --- | --- |
| ETC | ATPsynB (C.V) | RNAiA | ↓ FS (↑ SD) | Yes |
|  |  | RNAiB | ↓ FS (↑ SD) | Yes |
|  | ATPsynβ (C.V) | RNAiA | ↓ FS (↑ SD) | Yes |
|  | ATPsynγ (C.V) | RNAiA | ↓ FS (↑ SD) | Yes |
|  | ATPsynGL (C.V) | RNAiA | ⊘ | ⊘ |
|  | blw (C.V) | RNAiA | ↓ FS (↑ SD) | Yes |
|  | ATPsynC (C.V) | RNAiB | ↓ FS (↑ SD) | Yes |
|  | mtND2 (C.I) | RNAiB | ⊘ | ⊘ |
|  |  | RNAiC | ↓ FS (↓ DD) | ⊘ |
|  | ND-75 (C.I) | RNAiB | ↓ FS (↑ SD) | ⊘ |
|  | SdhC (C.II) | RNAiC | ↓ FS (↓ DD) | ⊘ |
|  | Cyt-c1 (C.III) | RNAiA | ↓ FS (↑ SD) | ⊘ |
|  |  | RNAiB | ↓ FS (↑ SD) | ⊘ |
|  | mtCo1 (C.IV) | RNAiB | ⊘ | ⊘ |
|  |  | RNAiC | ↓ FS (↓ DD) | ⊘ |
| Fission fusion | Drp1 | RNAiA | ⊘ | ⊘ |
|  | Marf | RNAiA | ↓ FS (↑ SD) | ⊘ |
|  |  | RNAiB | ↓ FS (↑ SD) | ⊘ |
|  | Opa1 | RNAiB | ↓ FS (↑ SD) | ⊘ |
|  |  | RNAiC | ↓ FS (↓ DD) | ⊘ |
| Cellular recycling | Park | RNAiA | ⊘ | ⊘ |
|  |  | RNAiB | ⊘ | ⊘ |
|  | Diap | RNAiC | ⊘ | ⊘ |
| Mitochondrial transporter | Rim2 | RNAiA | ⊘ | ⊘ |
|  |  | RNAiB | ⊘ | ⊘ |
| Mitochondria translation initiation factor | mIF2 | RNAiC | ↓ FS (↓ DD) | ⊘ |
| Heat shock protein chaperones 60 | Hsp60A | RNAiC | ⊘ | ⊘ |
| Mitochondrial biogenesis | Srl | RNAiB | ⊘ | ⊘ |
|  |  | RNAiC | ⊘ | ⊘ |
| Maintains cardiolipin | Taz | RNAiB | ⊘ | ⊘ |

**Supplementary Table 3:**
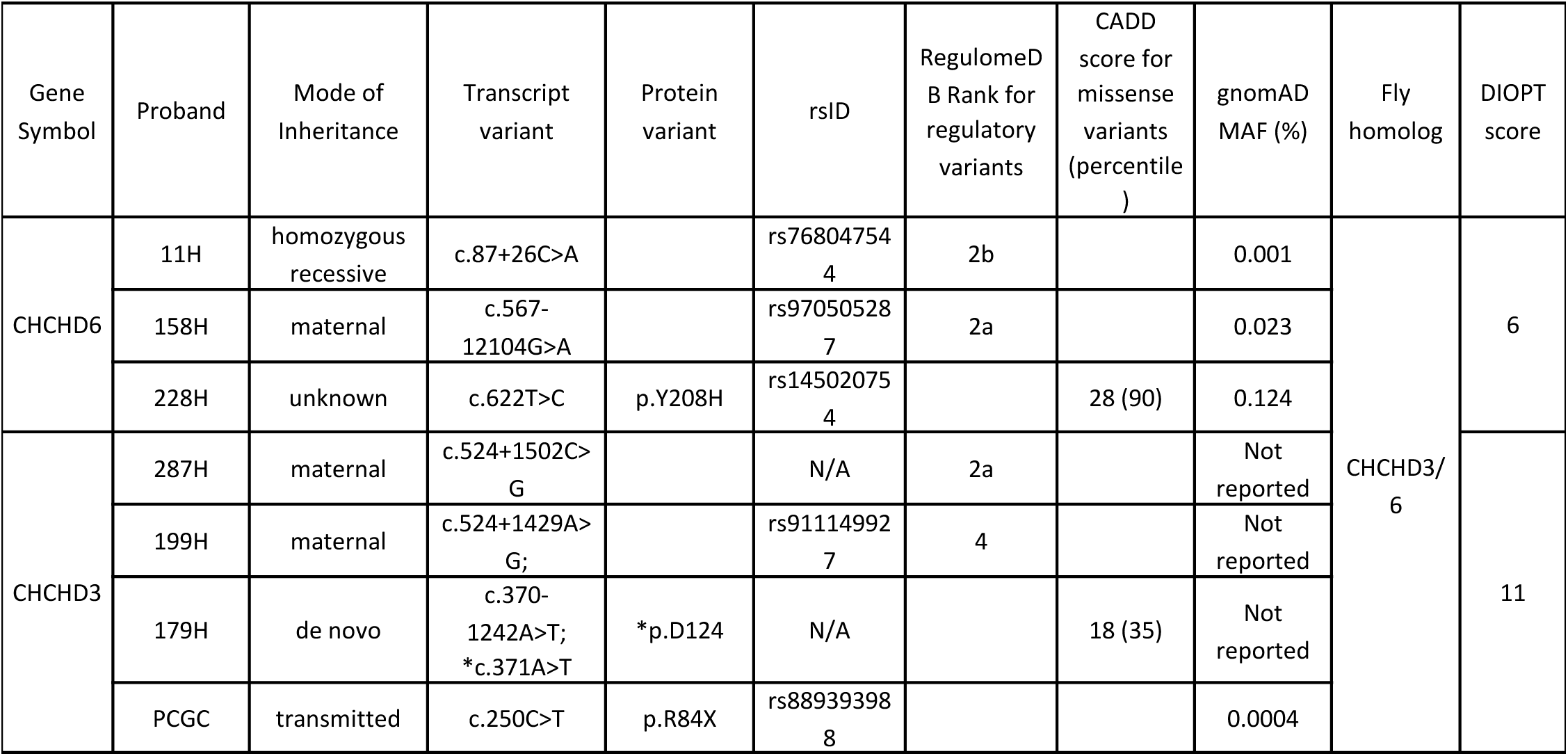
CHCHD3 and CHCHD6 MICOS variants in HLHS probands. Rare, predicted-damaging variants in CHCHD3 and CHCHD6 were identified in 6 of 183 HLHS probands in the Mayo Clinic cohort and the Pediatric Cardiac Genomics Consortium (PCGC). Transcript and relevant protein variants are listed. Corresponding *Drosophila* orthologs and orthology scores (DIOPT) are additionally listed (Hu *et al*., 2011). Note that probands 87H and 363H both harbor *APOO* and *SAM50* variants.

**Supplementary Table 4:**
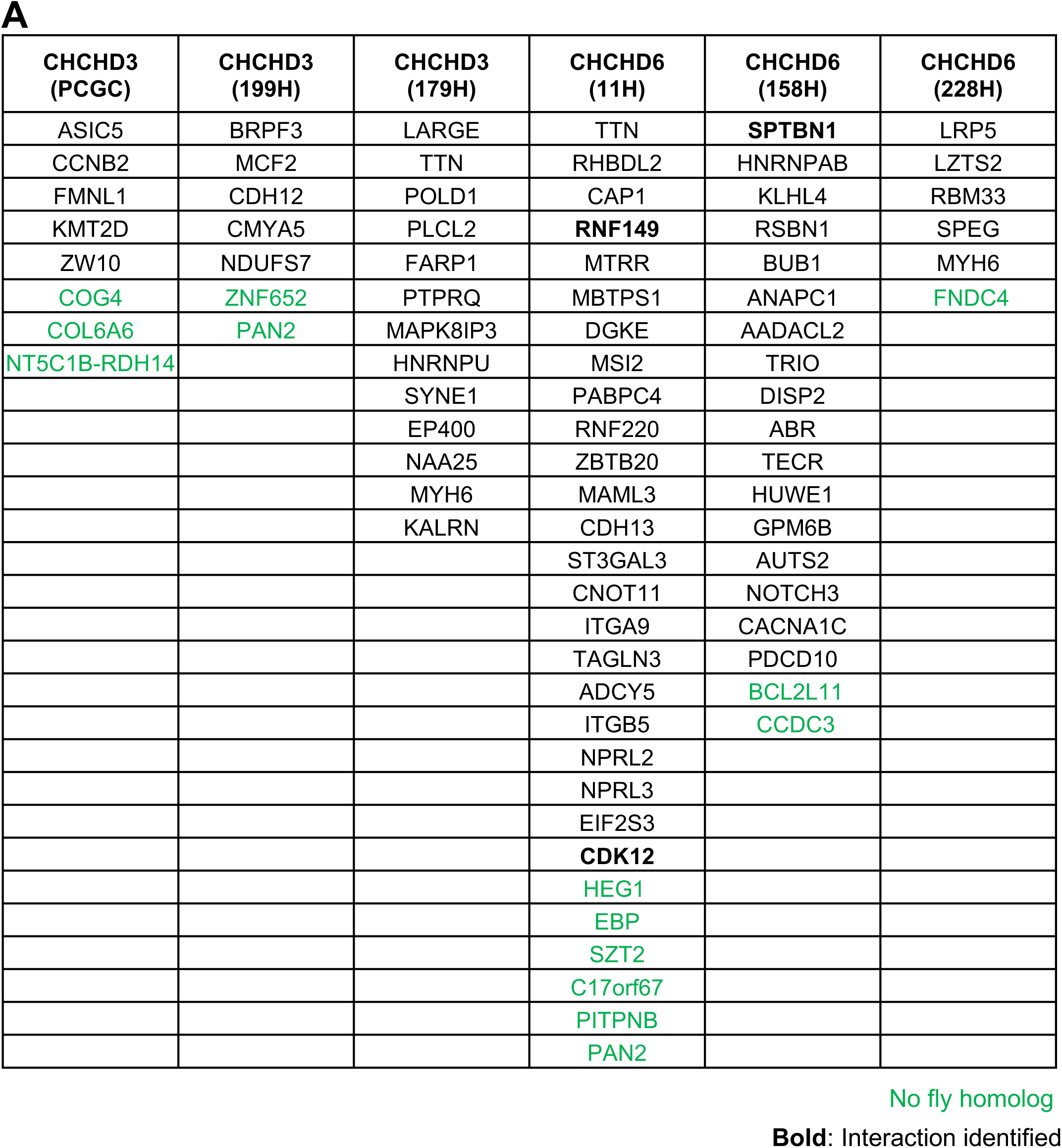
Candidate genes from HLHS probands harboring *CHCHD3* or *CHCHD6* variants. The majority of HLHS candidate genes has a *Drosophila* ortholog (genes without ortholog highlighted in green).

## Notes

### Competing Interest Statement

The authors have declared no competing interest.

### Author Declarations

Institutional Review Board of Mayo Clinic gave ethical approval for this work.

